# Regional and Temporal Patterns of COVID-19 Vaccine Misinformation in Sub-Saharan Africa

**DOI:** 10.1101/2025.08.19.25334033

**Authors:** Reyhan Jamalova, Devansh Jain, Sunny Rai, Juhi Mittal, Anietie Andy, Alison M. Buttenheim, Sharath Chandra Guntuku

## Abstract

**Background:** Despite a global rollout of COVID-19 vaccines, Sub-Saharan Africa lagged behind other regions in vaccination coverage, driven primarily by inequitable vaccine distribution and affordability barriers. Compounding this gap, COVID-19 vaccine hesitancy, fueled by concerns about side effects, distrust in institutions, and misinformation circulating on social media platforms like Twitter, further suppressed uptake even where vaccines were available.

**Methods:** We collected, preprocessed, and geolocated 6,305,628 tweets related to COVID-19 vaccination from Sub-Saharan Africa, and used a validated LLM-based classification framework to identify 1,312,969 tweets containing misinformation. We characterized the relationship between vaccine topics and misinformation prevalence, its variation over time, and its distribution across income-based and sub-regional country clusters.

**Results:** Misinformation-associated discourse centered on vaccine safety concerns, institutional distrust, and opposition to mandates. Sub-regional clustering revealed far more discourse variation (87 of 100 topics) than income-based clustering (7 of 100 topics), a pattern robust to excluding each region’s or cluster’s highest-volume country, with misinformation prevalence highest in Southern Africa and higher-income countries. Eastern and Western Africa’s discourse was largely implementation-focused, while Southern and Central Africa’s was more skepticism- and conspiracy-driven. Misinformation prevalence rose sharply around the introduction of vaccine mandates in late 2021 and remained elevated despite declining discourse volume.

**Conclusions:** These findings suggest that counter-misinformation strategies in Sub-Saharan Africa should be regionally tailored rather than uniform, and that public health communication should extend well beyond the rollout period, particularly after contentious policy decisions such as mandates, when misinformation appears to become more entrenched.

## Introduction

On May 5, 2023, the World Health Organization (WHO) declared an end to COVID-19 as a global health emergency. The unprecedented global effort undertaken to develop and adopt highly effective vaccines against COVID-19 played a crucial role in the slowdown of the pandemic [1]. However, as of December 31, 2023, only 39% of Africa’s population had received at least one dose of a COVID-19 vaccine, compared with 88% in the Western Pacific, 81% in the Americas, 77% in South-East Asia, and 69% in Europe [2]. Two primary reasons for this disparity are global vaccine distribution inequity [3] and vaccine hesitancy [4].

Within Sub-Saharan Africa (SSA) [46], vaccine hesitancy and under-vaccination are shaped by a complex interplay of demographic and structural barriers, institutional trust, and the circulation of vaccine-related misinformation [5, 6, 8, 10–12]. Prior research shows that being male, older (especially over 60), and more educated is often associated with greater COVID-19 vaccine acceptance [6, 11], though, one study [5] conducting national phone surveys found that in some countries, like Burkina Faso, Ethiopia, Malawi, and Nigeria, those with more years of education are significantly less willing to be vaccinated. The same study also identifies pockets of hesitancy among wealthier and urban populations. For routine childhood immunization, structural factors such as rurality, distance to health facilities, financial hardship, maternal autonomy, and lack of partner support are the most significant barriers to uptake [8, 10]. Importantly, even when vaccines are provided free of charge, hidden costs like transportation to clinics, childcare responsibilities, and the opportunity cost of missing work remain substantial deterrents for many households in resource-constrained settings [8, 11].

Institutional trust has been another consistent predictor of vaccination intention across the African continent. Trust in governments and ministries of health, as well as perceived social norms (believing that others intend to vaccinate), strongly predict vaccine uptake [12]. In many African contexts, this sense of collective responsibility resonates with the philosophy of Ubuntu (“I am because you are”), which emphasizes communal well-being and mutual interdependence [4]. Conversely, distrust in pharmaceutical companies rooted in colonial histories of medical exploitation fuels narratives that Africans are being used as experimental subjects, contributing to vaccine hesitancy [11, 12]. Concerns about vaccine safety and side effects remain among the most frequently cited reasons for refusal in national surveys [5, 7, 11]. Notably, healthcare workers, who serve as both clinical providers and influential role models in shaping public opinion, have exhibited substantial hesitancy themselves, with pooled estimates in SSA reaching approximately 46%, potentially undermining broader public confidence [7]. These dynamics suggest that vaccine confidence in SSA is shaped not only by access constraints but also by broader systems of trust and collective belief.

Online misinformation across social media platforms further amplifies these existing distrust and safety concerns [11, 13]. While social media platforms such as X (formerly Twitter) generally disseminate news, viewpoints, knowledge, and public health guidance around COVID-19 and vaccine-related issues, misinformation and intentional disinformation about COVID-19 and vaccines also circulate [11, 14, 15]. WHO coined the term “infodemic” to describe the overabundance of information (both accurate and false) during a disease outbreak [4, 19, 20]. Prior research demonstrates that exposure to vaccine-related misinformation on social media is associated with declining confidence and increased vaccine reluctance, as platforms amplify narratives about vaccine harm, infertility, or deliberate injury that accelerate distrust even among educated populations [4, 6, 11, 12, 16–18].

Given this central role in shaping public discourse, platforms such as X (formerly Twitter) have become important tools for studying vaccine-related attitudes and misinformation. X has been leveraged to measure changes in mental health [21], identify misinformation [22], study psychosocial effects [23], and uncover emerging symptoms [24]. According to a review of 45 studies about COVID-19 vaccine misinformation on social media platforms [25], X [26–28] was the most studied platform. These platforms provide rich, in-the-moment self-disclosed information on the attitudes towards and concerns about vaccination across communities. Such data can also complement large-scale surveys, which are often challenging to field quickly in resource-constrained contexts in multiple languages. However, the vast majority of research on social media and misinformation generally, including studies specific to X and COVID-19 vaccines, has been conducted using data from the United States and Europe [29, 30]. Less studied is the prevalence and characterization of online misinformation among non-WEIRD (Western, Educated, Industrialized, Rich, Democratic) countries.

In this study, we address this gap by 1) collecting a large-scale dataset of COVID-19 vaccination-related tweets from 44 Sub-Saharan African countries (SSA) between December 2020 and September 2022, 2) implementing a validated large language model-based content classification framework to detect misinformation-related tweets, and 3) characterizing the temporal and geospatial landscape of COVID-19 vaccine misinformation across SSA income clusters and sub-regions [47]. We identify statistically significant patterning of COVID-19 vaccine topics and misinformation prevalence across clusters.

## Materials and methods

### Data Collection and Pre-processing

In December 2020, we launched a data puller using the Python package TwitterMySQL [48] to collect tweets matching at least one of the following keywords, following previous work [29]: *vaccine, vaccination, moderna, pfizer, #antivax, #CashingInOnCovid, #MyBodyMyChoice, #Vax* via the official Twitter Application Programming Interface (API) available at the time. All data collection and subsequent analyses were conducted in accordance with Twitter’s Terms of Service and developer policies in effect at the time of data collection. The data puller continuously collected tweets using the Stream API until September 30, 2022, totaling **587,152,450** tweets globally. While this keyword set did not capture all locally specific vaccine terminology, several terms that referred to globally recognized entities (e.g., *moderna, pfizer, #Vax*) captured non-English vaccine-related discussions as well. Although English accounted for approximately 90% of all tweets from Sub-Saharan Africa, the remaining tweets spanned 60 identifiable languages (see Table S2).

### Geolocation

Mapping tweets to geographical locations around the world is non-trivial [31], since only a small fraction of tweets have geolocation coordinates that can be mapped directly. Thus, following prior work [12], we instead relied on parsing the free-response self-report location field that accompanies a tweet [49]. This field sometimes contains an administration level just by itself (e.g., “Johannesburg”, “Gauteng”, “South Africa”), or multiple administration levels together (e.g., “Johannesburg, Gauteng”, “Gauteng, South Africa”), or even non-identifiable phrases (e.g., “Who cares man”).

We began by tokenizing strings with “,” as the delimiter, based on the assumption that different administration levels in a location string are separated by commas, such that the number of “,” in a location string represents the number of administration levels mentioned. Given the number of administration levels, we then attempted to match the location string to the mapping dictionaries detailed in subsection S2, starting from the highest administration level. We also utilized substring lookup for location strings having more than one administration level. Specifically, if the entire location string did not yield a valid match in the corresponding mapping dictionaries, we removed the lowest administration level from the string and repeated the mapping process. This was helpful when the lowest administration level in a string (usually a city) was absent in the mapping dictionaries, but the string contained information about higher administration levels that could still be used to successfully identify its location, albeit with a loss of granularity. Before applying this algorithm to the full dataset, we did a sanity check by randomly sampling 100 geolocated tweets and manually verifying whether the mapped country corresponded to the self-reported location. The comma-based tokenization with hierarchical fallback correctly mapped 95% of sampled tweets. We also decided to exclude the Democratic Republic of Congo (DRC) from geolocation due to persistent ambiguity in distinguishing “Congo” from “DRC” in self-reported location fields, as well as substantial variation in how users referenced this country, making it difficult to map them appropriately using our heuristic-based geolocation algorithm.

We then applied a geolocation algorithm and assessed the country-level geolocation accuracy by randomly sampling up to 3,000 tweets per each SSA country and manually verifying whether the mapped country corresponded to the self-reported location. 1,696,693 entries identified as false positives during this audit were eliminated from the geolocated dataset. With this false-positive removal step, 91,804,465 of 587,152,450 tweets (15.6%) were mapped to 59 countries worldwide. Of these, 46 were Sub-Saharan African (SSA) countries, comprising 6,310,950 tweets. No tweets were geolocated to SSA countries Eswatini or Namibia.

Next, we removed Sudan and South Sudan from the analytic dataset because missing GDP per capita data precluded their assignment to an income cluster. To ensure consistency across analyses, all subsequent income-cluster and sub-regional analyses were conducted on the same set of countries. Following these exclusions, the final Sub-Saharan Africa dataset consisted of **6,305,628** tweets (D_Full_) posted by **572,055** users. A deduplicated version (D_Dedup_), generated by removing duplicate cleaned tweets, contained **2,909,260** tweets (see subsection S1 for details) and was used for topic modeling and misinformation classification. Language- and country-level coverage of D_Full_ and D_Dedup_ is reported in Table S2 and Table S3, respectively.

### Topic Modeling

We created a set of topics using Latent Dirichlet Allocation (LDA; [33]), a probabilistic generative model that assumes tweets are generated by a combination of topics (which are latent variables) and that topics are distributions over words. We first removed the top 100 most frequent words in the dataset, along with a manually curated list of stopwords (see subsection S3). Using the DLATK library’s [34] interface for the MALLET implementation of LDA [35], we generated 50, 100, and 200 topics, with an alpha level of 5, the default MALLET configuration [50]. Following [36], we quantitatively assessed the quality of topics in each set by computing three topic coherence measures and topic uniqueness, and examining their behavior using an elbow plot (see Figure S1) [37]. We also reviewed the top words of topics within each topic set for a qualitative check of interpretability. Using both methods, we reached the consensus that the 100-topic model was optimal. Finally, we obtained the probability distribution for each tweet across all 100 topics.

### Cluster Identification

To examine trends in vaccine discourse across Sub-Saharan Africa, we grouped countries into broader socioeconomic clusters rather than analyzing individual countries separately, as some countries contributed insufficient data for reliable country-level analyses. We derived these clusters using agglomerative hierarchical clustering based on GDP per capita [40]. Using a dendrogram, we selected 3 clusters, which broadly corresponded to low-income, lower-middle-income, and high-/upper-middle-income country groups.

In addition, we grouped countries into the four African sub-regions—Central, Eastern, Southern, and Western Africa—to examine broader geographic patterns in vaccine discourse. Table S3 presents the country assignments for both income clusters and African sub-regions. To facilitate interpretation of these groupings, Table S4 and Table S5 report country-level COVID-19 case rates, death rates [38], vaccination rates [39], Corruption Perceptions Index (CPI), Democracy Index (DI), and cellular phone subscription rates [40], as well as the distributions of tweets, users, retweets, and countries within each grouping.

### Misinformation Classification

We define misinformation as the presence of an **unverified claim**. Tweets were labeled using three categories: **Yes** (contains an unverified claim), **No** (does not contain an unverified claim), and **Unsure** (insufficient context to determine verification status).

We developed a content-based classification framework using prompt engineering with GPT-4o-mini (final prompt in subsection S2). To evaluate performance, we manually annotated a random sample of 100 tweets and compared these labels to model outputs.

The final prompt achieved **Weighted Precision = 0.943**, **Weighted Recall = 0.920**, and **Weighted F1 = 0.927**. For the misinformation (*Yes*) class specifically, performance was **Precision = 0.74**, **Recall = 0.93**, and **F1 = 0.82**, indicating strong recall with moderate precision in identifying unverified claims. Applying this validated classifier to the deduplicated dataset resulted in the identification of **599,057 tweets** containing misinformation, accounting for approximately **21% of all tweets** in *D_Dedup_*.

In downstream analyses, we treated tweets classified as “Unsure” as equivalent to “No” (i.e., coded as 0), on the grounds that only tweets with a clear, confident classification of misinformation (“Yes”) were counted as containing an unverified claim.

### Differential Language Analysis

We performed a differential language analysis to identify statistically significant correlations between the topics and different outcomes. We extracted the quarterly prevalence of topics across tweets aggregated to the second-highest administration level (aka, province/state) to assess trends over time. Topics were then used as input in a logistic regression model with dummy variables for each quarter as the outcomes for temporal analyses. For the country income clusters and African sub-regions, we aggregated the probability distribution of tweet-topics to the second-highest administration level and used them as input in a logistic regression model with dummy variables for each of the cluster sets as the outcomes. For the misinformation outcome, we conducted a tweet-level analysis with topics as independent variables in a logistic regression model to predict whether a tweet contained misinformation. Based on conventional linguistic analysis, we used a p-value of *<* 0.05 to identify significant linguistic markers, and all p-values were corrected for the false discovery rate during multiple hypothesis testing using the Benjamini-Hochberg correction.

## Results

### Country Clusters

Three clusters emerged from the income and four from African sub-regional clustering analyses (Figure 1). Cluster 1, representing low-income countries, comprises 24 countries such as Benin, Niger, Somalia, Zambia, etc.; Cluster 2 (lower-middle-income countries) contains 14 countries such as Kenya, Nigeria, Senegal, etc.; and Cluster 3 (High/Upper-middle-income countries) has 6 countries, i.e. Botswana, Equatorial Guinea, Gabon, Mauritius, Seychelles, South Africa. For African sub-regions, we used the UN M49 classification, resulting in 17 countries in Eastern Africa, 16 in Western Africa, 8 in Central Africa, and 3 in Southern Africa.

**Fig 1.**
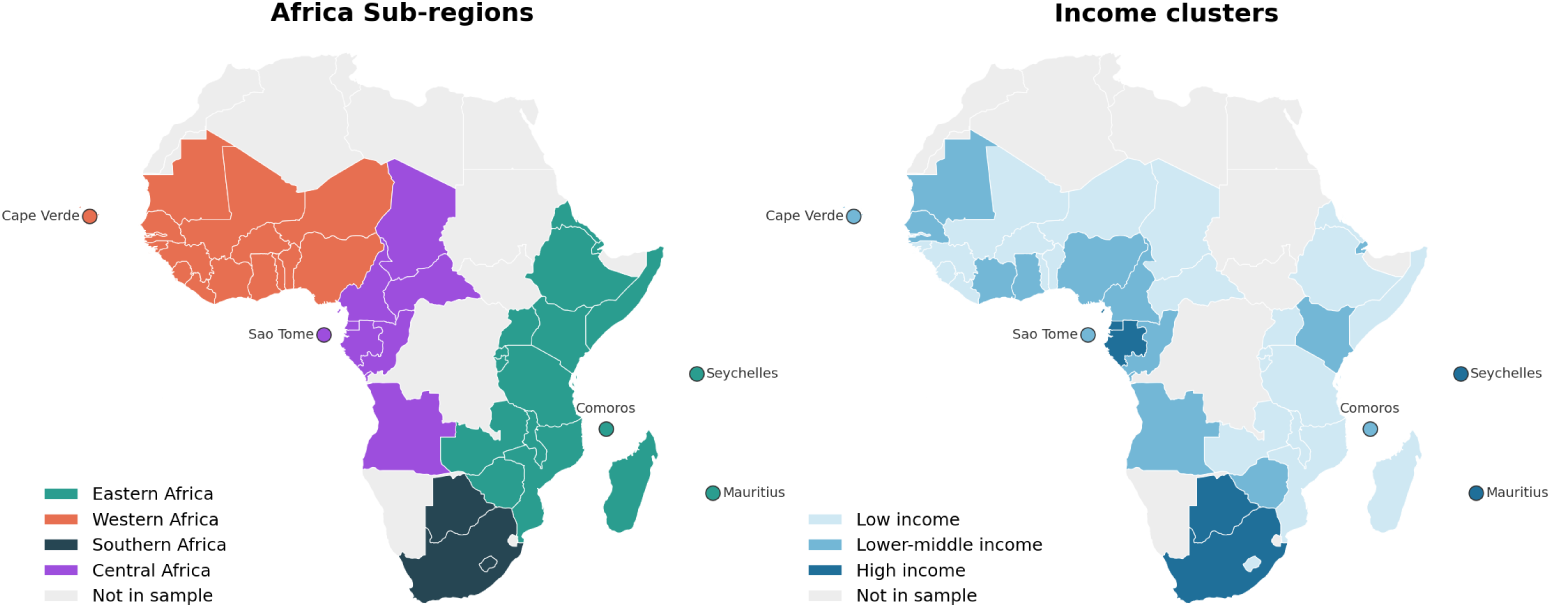
Choropleth maps showing the African sub-regions (UN M49 classification) and income-cluster classifications for the 44 SSA countries included in the analysis. The left panel shows the four African sub-regions: Eastern Africa, Western Africa, Central Africa, and Southern Africa. The right panel shows the income-cluster groupings, consisting of low-income, lower-middle-income, and high-/upper-middle-income countries derived through agglomerative hierarchical clustering. Countries shown in gray were not included in the study.

### Misinformation

Out of 6,305,628 tweets in *D_F ull_*, our misinformation classifier predicted 1,312,969 tweets (20.8%) to contain misinformation. The misinformation rate (number of misinformation posts/number of total posts) varied by both regional and income clusters, with Southern Africa and the high-/upper-middle-income countries exhibiting the highest prevalence of misinformation (Table 1). Topics most strongly associated with misinformation included vaccine safety concerns, including alleged adverse events, deaths, blood clots, myocarditis, and claims that authorities or health organizations had concealed evidence of vaccine harms. Misinformation was also characterized by distrust of governments, pharmaceutical companies, and public health institutions, alongside skepticism about vaccine testing and effectiveness, the sharing of anecdotal accounts of injury or death, and opposition to vaccine mandates, passports, and other pandemic-related restrictions (Table 2).

**Table 1.** Misinformation prevalence across African sub-regions and income clusters. Counts and percentages of tweets classified as misinformation are shown for each sub-region and income cluster. Southern Africa and the high-/upper-middle-income cluster exhibited the highest prevalence of misinformation.

| Misinformation Rates across African Sub-regions |  |  |  |
| --- | --- | --- | --- |
| Sub-region | #Tweets | #Misinfo | Misinfo (%) |
| Central Africa | 123,727 | 26,069 | 21.10% |
| Eastern Africa | 1,357,454 | 185,065 | 13.60% |
| Southern Africa | 3,494,611 | 895,365 | 25.60% |
| Western Africa | 1,329,836 | 206,470 | 15.50% |
| Misinformation Rates across Income Clusters |  |  |  |
| Income Cluster | #Tweets | #Misinfo | Misinfo (%) |
| Low income | 643,636 | 97,583 | 15.20% |
| Lower-middle income | 2,153,417 | 315,959 | 14.70% |
| High/Upper-middle income | 3,508,575 | 899,427 | 25.6% |

**Table 2.** Topics associated with misinformation. The top 10 topics associated with misinformation are shown, along with representative top keywords, Pearson correlation coefficients (*r*), and 95% confidence intervals. These are primarily focused on vaccine safety concerns (e.g., adverse events, myocarditis, clinical trials), distrust of pharmaceutical companies and institutions, and opposition to mandates and vaccine passports.

| Topic | Top Keywords | $r$ | CI |
| --- | --- | --- | --- |
| 91 | data, adverse, report, deaths, events, release, safety, released, cdc, documents | 0.149 | 0.148, 0.15 |
| 96 | blood, heart, clots, death, died, rare, cause, myocarditis, receiving, due | 0.146 | 0.146, 0.147 |
| 16 | these, them, into, stop, fear, experimental, pushing, lies, keep, truth | 0.141 | 0.141, 0.142 |
| 7 | trials, trial, clinical, tested, safe, testing, used, safety, test, ivermectin | 0.130 | 0.129, 0.131 |
| 74 | were, would, said, did, had, told, thought, didn't, never, knew | 0.102 | 0.101, 0.102 |
| 56 | big, pharma, companies, money, pharmaceutical, company, business, making, profit, profits | 0.099 | 0.099, 0.1 |
| 38 | don't, want, them, know, dont, ppl, let, trust, vaccinate, die | 0.098 | 0.098, 0.099 |
| 37 | she, her, got, his, friend, died, family, mom, told, old | 0.092 | 0.092, 0.093 |
| 43 | passports, mandates, passport, freedom, mandatory, lockdowns, control, society, vote, restrictions | 0.091 | 0.09, 0.091 |
| 65 | don't, it's, know, doesn't, because, work, even, want, can't, that's | 0.089 | 0.088, 0.09 |

Misinformation prevalence also fluctuated across the study period (2020 Q4-2022 Q3), ranging from 14.4% (2021 Q2) to 36.9% (2022 Q2) (Table 3). Rates were moderate during initial vaccine rollout (2020 Q4-2021 Q1: 19.1%-16.4%), declined slightly through mid-2021 (14.4%-18.7%), then rose steadily from late 2021 onward (23.7% in 2021 Q4), peaking at 32.9%-36.9% across 2022 Q1-Q3. This upward trend in misinformation prevalence coincided with a substantial decline in overall tweet volume, from a peak of 1,559,460 tweets in 2021 Q3 to 167,586 tweets by 2022 Q3.

**Table 3.**
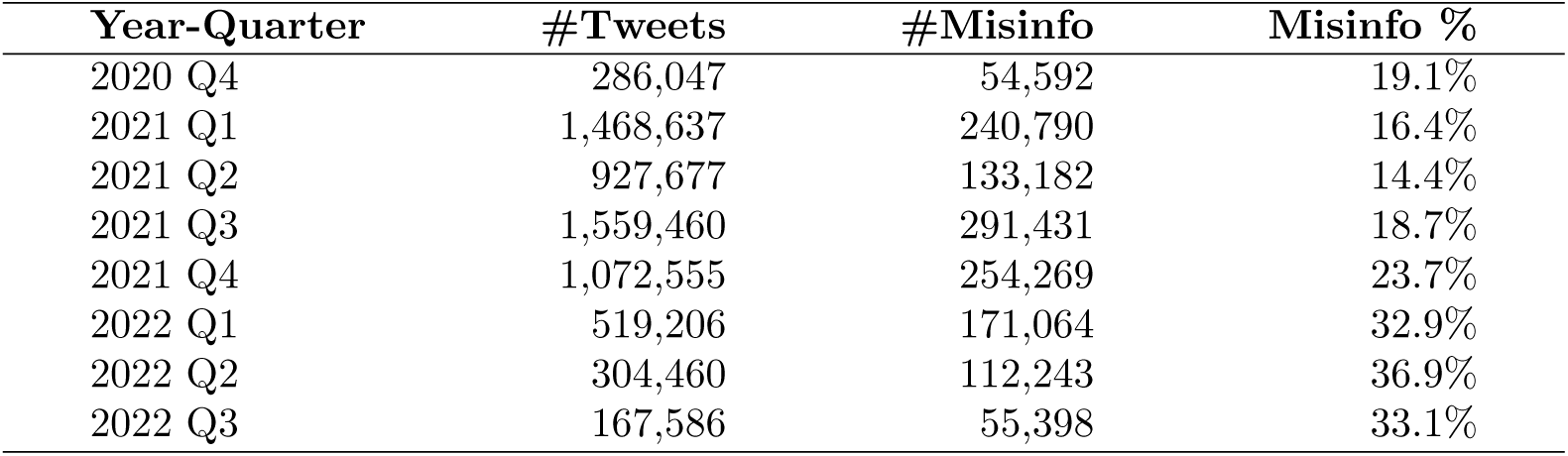
Quarterly tweet volume and misinformation prevalence in vaccine-related discourse across Sub-Saharan Africa, 2020 Q4-2022 Q3. #Tweets and #Misinfo denote the total number of tweets and the number classified as containing misinformation, respectively, for each quarter; Misinfo % represents the proportion of tweets classified as misinformation (#Misinfo / #Tweets). Misinformation prevalence nearly doubled over the study period, rising from 19.1% in 2020 Q4 to a peak of 36.9% in 2022 Q2.

| Year-Quarter | #Tweets | #Misinfo | Misinfo % |
| --- | --- | --- | --- |
| 2020 Q4 | 286,047 | 54,592 | 19.1% |
| 2021 Q1 | 1,468,637 | 240,790 | 16.4% |
| 2021 Q2 | 927,677 | 133,182 | 14.4% |
| 2021 Q3 | 1,559,460 | 291,431 | 18.7% |
| 2021 Q4 | 1,072,555 | 254,269 | 23.7% |
| 2022 Q1 | 519,206 | 171,064 | 32.9% |
| 2022 Q2 | 304,460 | 112,243 | 36.9% |
| 2022 Q3 | 167,586 | 55,398 | 33.1% |

### Topic Analysis by African Sub-regions

Regional differences in vaccine discourse were substantial, with 87 of 100 topics significantly overrepresented in at least one African sub-region (Figure 2). **Eastern Africa** was characterized by discussions of vaccine rollout, access, community engagement, vaccination campaigns, and vaccine procurement, reflecting a largely implementation-focused discourse. **Western Africa** was dominated by Nigeria-centered conversations, including government announcements, news sharing, and public discussion of vaccination policies; notably, none of Western Africa’s three significant topics were misinformation-associated. **Southern Africa** showed greater emphasis on vaccine skepticism and misinformation-related narratives, including distrust of authorities, concerns about vaccine safety, adverse-event reports, and opposition to vaccination efforts. **Central Africa** was distinguished by discussions in French and conspiracy-related narratives, including topics explicitly centered on conspiracy theories and misinformation. Overall, misinformation-associated topics were concentrated primarily in Southern and Central Africa, whereas Eastern and Western Africa were more strongly characterized by vaccine rollout and information-sharing discussions.

**Fig 2.**
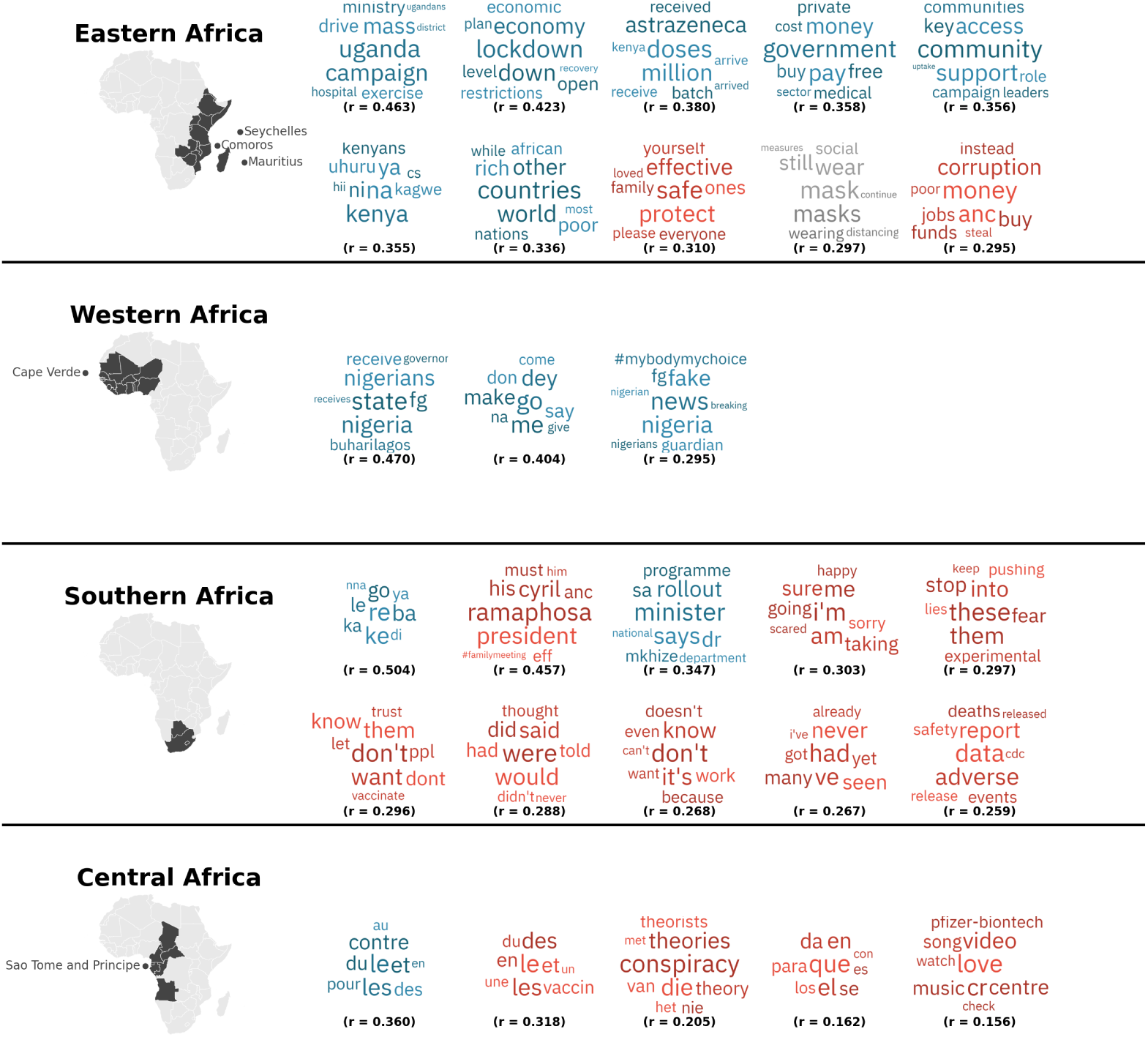
Regional variation in COVID-19 vaccination discourse across Sub-Saharan Africa. For each African sub-region, the figure shows up to ten topics most strongly overrepresented relative to the other regions. Topic cloud colors denote their relationship with misinformation discourse: **red** indicates topics positively associated with misinformation, **blue** indicates topics negatively associated with misinformation, and **gray** indicates no significant association. The correlation coefficients (*r*) shown below each topic cloud represent the strength of association between the topic and the focal region. Only topics significantly associated with a region after Benjamini-Hochberg correction (*p <* 0.05) are displayed. Eastern Africa is characterized primarily by vaccine rollout, access, and community-support discussions; Western Africa by information-sharing discourse centered largely on Nigeria; and Southern and Central Africa by a greater concentration of misinformation-associated topics, including distrust, adverse-event, and conspiracy-related narratives.

Tweet volume was highly concentrated within each sub-region, with South Africa, Nigeria, Kenya, and Cameroon accounting for 94.0%, 65.7%, 48.1%, and 71.2% of their respective regions’ tweets. Given this disproportionate distribution, we conducted a robustness check to assess whether the regional characterizations reported above depended on each region’s single highest-volume country, rather than reflecting discourse patterns distributed more broadly across each region’s constituent countries (see Figure S3). Sub-regional clustering remained associated with substantial topic variation after this exclusion, with 80 of 100 topics remaining significantly overrepresented in at least one sub-region. Eastern Africa’s implementation-focused characterization was largely retained, with rollout, dose delivery, and booster-related themes continuing to dominate its top topics. Southern Africa’s skepticism- and distrust-oriented tone also persisted essentially unchanged, with government-performance criticism, fear, and distrust themes remaining prominent after excluding South Africa. Western Africa and Central Africa, which had fewer significantly overrepresented topics in the full sample (3 and 6 of 100, respectively), showed more variable topic composition under exclusion: Central Africa’s conspiracy-theory-specific topic, present in the full-sample analysis, did not persist after excluding Cameroon, suggesting this narrative element may be disproportionately attributable to Cameroon rather than broadly characteristic of Central Africa; conversely, Western Africa’s expanded set retained the persisting non-misinformation Pidgin-conversational topic alongside three additional French-language topics.

### Topic Analysis by Income Clusters

We also identified topics associated with different income clusters. Out of 100 topics, only seven were significantly overrepresented after Benjamini-Hochberg correction (see Figure S2), suggesting limited variation in vaccine discourse by income level. Topics overrepresented in the high-/upper-middle-income cluster primarily reflected South African political discourse and local-language discussions, whereas those overrepresented in the lower-middle-income cluster focused on Nigerian politics and governance, and healthcare and disease-related discussions (e.g., HIV, cancer, HPV). No topics were significantly overrepresented in the low-income cluster.

As a robustness check, we repeated this analysis after excluding South Africa and Nigeria, the highest-volume contributors to the high-/upper-middle-income and lower-middle-income clusters, respectively; the low-income cluster was not included in this check, as it already showed no significantly overrepresented topics in the full-sample analysis. None of the seven originally significant topics remained overrepresented after exclusion, and no alternative topics emerged (0 of 100), indicating that the income-cluster effect was entirely attributable to these two countries rather than reflecting broader income-level patterns.

### Topic Analysis Over Time

Topic prevalence varied substantially over time, with 85 of 100 topics significantly associated with at least one calendar quarter (Figure 3). The introduction and expansion of COVID-19 vaccination campaigns across Sub-Saharan Africa coincided with marked shifts in both the volume and content of vaccine-related discourse on X, as well as in the prevalence of misinformation (Table 3).

**Fig 3.**
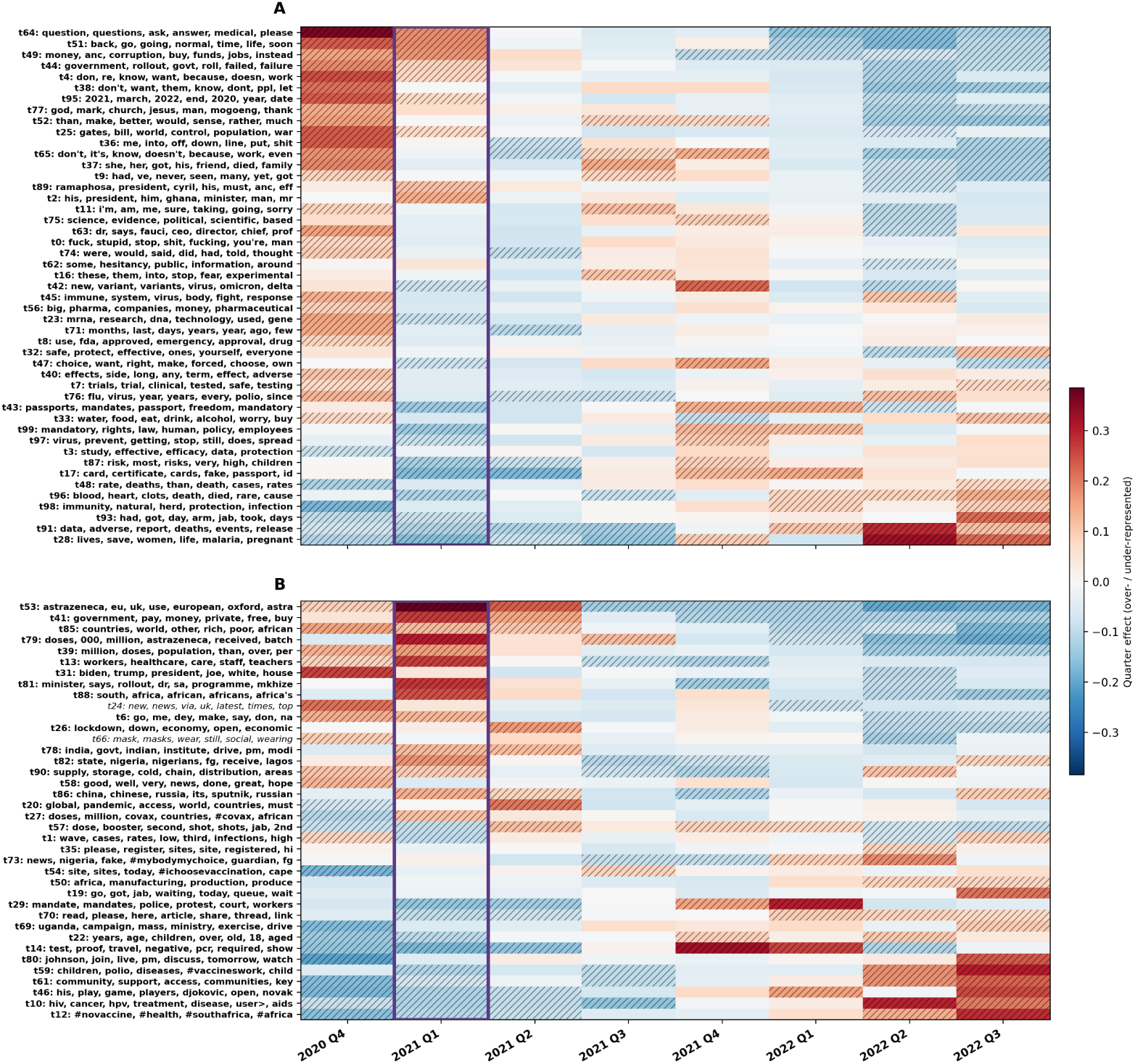
Temporal dynamics of vaccine-related discussion topics and COVID-19 vaccine availability across Sub-Saharan Africa. The heatmap shows quarter-specific over- and under-representation of vaccine-related topics from 2020 Q4 to 2022 Q3. Only topics that were significantly associated with at least one quarter are shown. **Red** cells indicate topics discussed more frequently than expected in a given quarter, while **blue** cells indicate lower-than-expected discussion. Hatched cells denote topics significantly associated with that quarter after Benjamini-Hochberg correction. Topic labels are shown using their most representative keywords; topics positively associated with misinformation are shown in panel A, whereas topics negatively associated with misinformation are shown in panel B. Topics written in italic do not have significant associations with misinformation. The purple outline highlights 2021 Q1, when most (31) Sub-Saharan African countries introduced COVID-19 vaccines. Cameroon, Comoros, Madagascar, Burkina Faso, Chad, Central African Republic, Guinea, Guinea-Bissau, Liberia, and Zambia started official vaccination campaigns in 2021 Q2, Tanzania in 2021 Q3, and Burundi in 2021 Q4. Eritrea did not introduce COVID-19 vaccination during the study period. There is a marked shift in vaccine-related discourse following vaccine introduction, with vaccine rollout, access, safety, and government trust concerns dominating early discussions and mandates, vaccine certificates, natural immunity, vaccine side-effects, and broader health topics becoming more prominent as vaccination campaigns progressed.

**2020 Q4.** Discourse prior to vaccine rollout was dominated by uncertainty: speculation about a return to normalcy, criticism of government handling of the pandemic, religious framing of the crisis, and conspiracy narratives involving global control and Bill Gates. These misinformation-associated topics coexisted with a substantial volume of non-misinformation discourse concerning vaccine cost and pricing, disparities between wealthy and low-income countries in vaccine access, vaccine distribution planning, and infection reporting.

**2021 Q1.** Tweet volume increased approximately 4.5-fold relative to the prior quarter, coinciding with the period in which most countries in the sample (31/44) first introduced vaccines. Despite this surge in discourse, misinformation prevalence remained comparatively low; residual uncertainty and commentary on a return to normalcy persisted, but the dominant topics concerned practical aspects of rollout — vaccine fees, dose availability, and logistics of official rollout campaigns.

**2021 Q2.** Overall discourse volume declined modestly, and misinformation prevalence dropped further, with no topic showing significant overrepresentation of misinformation content. Non-misinformation discourse remained high and diversified, encompassing dose and booster scheduling, geopolitical vaccine sourcing (Chinese and Russian-manufactured vaccines), country-specific rollout milestones (e.g., Nigeria’s receipt of vaccine shipments, South Africa’s rollout), vaccine supply and storage logistics, prioritization of healthcare workers and teachers, and AstraZeneca-specific discourse.

**2021 Q3.** A second surge in discourse volume occurred alongside a rise in misinformation prevalence. While practical topics (dose scheduling, vaccination site information) persisted, misinformation discourse increasingly centered on anecdotal reports of death following vaccination, claims that vaccines were experimental or insufficiently tested, and questioning of vaccine effectiveness.

**2021 Q4.** Overall discourse volume declined somewhat, while misinformation prevalence continued to increase. This quarter marked the emergence of several new thematic clusters, most notably the Omicron variant and the introduction of vaccine mandates and passports. Mandate-related discourse was bidirectional: alongside misinformation-associated framing around personal choice, bodily autonomy, and claims that vaccination did not prevent transmission or infection, non-misinformation discourse addressed vaccine documentation for travel and mandate implementation. Concern regarding pediatric vaccination risk also emerged during this period. The introduction of mandates appears to represent a distinct inflection point shaping discourse for the remainder of the study period.

**2022 Q1.** Overall discourse volume declined sharply, while misinformation prevalence rose by approximately nine percentage points relative to the prior quarter: the largest single-quarter increase observed. This quarter saw the emergence of adverse-event discourse specific to blood clots and cardiac events, alongside claims favoring natural immunity over vaccination. Mandate-related discourse persisted.

Notably, this quarter also marked the first appearance of organized counter-misinformation behavior, with users sharing corrective links and explicitly urging others to “read” and “share” accurate information.

**2022 Q2.** Discourse volume continued to contract while misinformation prevalence reached its peak. Misinformation topics continued to center on adverse-event narratives, alongside comparisons to flu and polio vaccines. Concurrently, non-misinformation discourse included HIV-related discussion, community support and access initiatives, and continued attention to pediatric vaccination programs.

**2022 Q3.** Overall discourse reached its lowest observed volume, and misinformation prevalence declined slightly from its Q2 peak but remained elevated. Misinformation themes overlapped substantially with the prior quarter (adverse effects, natural/herd immunity, disputes over death versus case reporting rates), while non-misinformation discourse again featured community support, vaccine efficacy affirmations, and HIV/cancer-related discussion.

## Discussion

In this analysis of tweets originating in Sub-Saharan Africa (SSA) regarding the COVID-19 vaccine, we identified significant variations in the prevalence of latent topics over time and across socio-spatial clusters of countries in the region, defined by income-based and geographic sub-regional groupings. Drawing on over 6.3 million tweets across 44 SSA countries, this represents, to our knowledge, **the largest and most geographically comprehensive study of COVID-19 vaccine discourse using social media data conducted in the region to date.** Temporal trends in topic prevalence mirrored phases of the pandemic and reflected the news cycle as well, particularly around the development, roll-out, and later mandating of vaccines.

Income-based clustering revealed limited variation in discourse; sub-regional clustering, by contrast, revealed substantial and distinct differences across geographic areas. While about 1 in 5 tweets in the full dataset contained misinformation, a rate consistent with prior estimates reported in the literature [41], this rate varied substantially over the socio-spatial clusters and over time (see Table 1 and Table 3).

Our analysis makes several unique contributions to our understanding of the role of misinformation in COVID-19 vaccine hesitancy in a region that ultimately had low population-level vaccine coverage [2]. First, while most published studies on COVID-19 vaccine hesitancy in Sub-Saharan Africa rely on self-reported attitudes, beliefs, and information-seeking behaviors collected via survey [5, 6, 12, 42, 43, 45] and are often limited to one or a handful of countries, our analysis is based on naturally occurring social media discourse spanning nearly the entire region, offering a scale and breadth of coverage not previously available. Second, by jointly examining temporal, regional, and income-based variation, we offer a more granular account of vaccine discourse than single-country [43–45] or sub-regional [5, 6, 10, 12] analyses have provided; these insights can enhance targeted and precise public communication. A third important contribution is the development and validation of a LLM-based classifier to detect misinformation-containing tweets at scale across this large, linguistically and geographically diverse dataset. This allows us to describe temporal trends in the progression of misinformation over a span of two years, from a low-misinformation period dominated by practical rollout discourse in early 2021, through a clear inflection point following the introduction of vaccine mandates in late 2021, to a sustained rise in misinformation prevalence that persisted even as overall discourse volume declined through 2022.

Of our two clustering approaches, sub-regional grouping revealed far more variation in discourse (87 of 100 topics) than income-based clustering did (7 of 100 topics). We interpret this partly as a data-availability artifact rather than a true absence of income-related variation: low-income countries, though comprising the majority of the region, also have the lowest rates of internet and social media penetration, likely muting their signal even in a dataset of this scale. This aligns with the broader pattern that greater connectivity, more typical of wealthier, urban populations, may itself be a channel of exposure to misinformation-generating discourse, while lower-connectivity populations remain comparatively voiceless in analyses like ours.

Regional grouping also revealed distinct discourse “profiles”: Eastern and Western Africa’s discourse remained largely implementation-focused (rollout, access, government announcements), while Southern and Central Africa’s discourse was more heavily marked by distrust and conspiracy narratives. This heterogeneity, visible only because our data span nearly all of SSA rather than a handful of countries, suggests a single regional counter-misinformation strategy is unlikely to be effective; regions with entrenched skepticism may need targeted trust-repair and multilingual pre-bunking.

This connectivity-driven concentration of voice operates not only between clusters, muting low-income countries entirely, but also within them: robustness checks excluding each cluster’s or region’s highest-volume country showed that the modest signal observed in the higher-income clusters was itself concentrated in a single dominant country. The income-cluster effect, already modest, was eliminated entirely after excluding South Africa and Nigeria (0 of 100 topics remained overrepresented), while the sub-regional effect persisted with only a modest reduction (80 of 100 topics), indicating that geography, rather than income, is the more robust axis of discourse variation in this dataset. At the same time, this sub-regional variation was itself shaped disproportionately by a small number of high-volume countries within each region, suggesting that the discourse “profiles” described above partly reflect the visibility of a few highly active countries rather than uniformly distributed discourse across all countries in a region.

Turning to temporal variation, discourse shifted markedly around the introduction of vaccine mandates and passports in 2021 Q4, which coincided with a clear inflection point in the remainder of the study period. Before this point, misinformation prevalence had been comparatively low and discourse was dominated by practical rollout topics; after this point, misinformation prevalence rose sharply and adverse-event and bodily-autonomy narratives became more prominent features of the discourse, persisting even as overall volume declined from its 2021 Q3 peak. This temporal association is consistent with several possible explanations, which our data cannot distinguish between: mandates may have directly prompted a shift in discourse; the timing may instead reflect a broader confluence of concurrent events (e.g., the Omicron variant’s emergence, declining global media attention to vaccination); or the pattern may partly reflect survivorship bias, as a shrinking, increasingly self-selected group of remaining tweeters came to dominate the discourse as overall volume declined. If mandates do carry a lasting discursive cost, this would suggest that public health messaging may need to extend well beyond the rollout period, particularly following contentious policy decisions, rather than being front-loaded to the initial campaign.

### Limitations

Our study does have some important **limitations**. First of all, the keyword-based query we used to collect tweets may not exhaustively capture all locally specific vaccine terminology used across Sub-Saharan contexts, which may result in underrepresentation of vaccine discourse expressed exclusively through local or non-standard terms. Second, our analyses rely on data from X, which introduces several layers of selection bias. The population represented in our dataset is limited to individuals who (i) have access to smartphones, (ii) have reliable internet connectivity, (iii) maintain profiles on X, (iv) are sufficiently active on the platform, and (v) choose to engage with COVID-19 vaccine-related content. Prior research from Nigeria [44, 45] suggests that platform usage may be highly non-representative of national populations, often skewed toward younger, urban, and male users. These factors imply that our findings may not reflect the broader population in Sub-Saharan Africa, particularly individuals in rural or lower-connectivity settings who are less likely to participate in online discourse.

Third, despite our dataset’s breadth, our cluster- and regional-level findings partly reflect a small number of high-volume countries: South Africa accounts for 93.6% of the high-/upper-middle-income cluster and 94.0% of the Southern Africa sub-region, and Nigeria accounts for 65.7% of the Western Africa sub-region. As reported above, robustness checks excluding these and other dominant countries confirmed that the income-cluster effect was entirely attributable to country-level rather than income-level differences, whereas the sub-regional effect was more robust, though some region-specific content did not persist. Relatedly, since our topic model was trained on the pooled corpus without weighting for country-level volume, the resulting topics themselves may already disproportionately reflect high-volume countries’ discourse, independent of the clustering scheme applied downstream — a limitation our robustness checks, which held the topic model fixed, cannot address.

We also did not perform user-level demographic inference; prior survey research from Nigeria [45] suggests that age and education shape how health information is trusted and shared online, implying that discourse likely varies by demographic subgroup in ways our analysis cannot capture. Large-scale demographic inference from social media in low-resource settings raises its own methodological and ethical challenges, including sparse user activity, limited labeled data, and misclassification risk [44], though future work could explore name-based matching, graph-based propagation, or culturally grounded validation approaches to address this gap.

Additionally, our misinformation detection model exhibits low precision (0.74), primarily attributed to the prevailing tone of fear in COVID-19-related tweets. This fear-inducing tone is often found in tweets containing misinformation, and developing a more nuanced model capable of contextualizing fear (such as distinguishing fear related to vaccines from fear related to infection) would lead to more precise predictions. More broadly, our reliance on an LLM-based classifier introduces the possibility of systematic bias inherited from the model’s training data and design, which a validation exercise on a relatively small sample of 100 tweets may not fully surface; larger and more diverse validation sets, alongside comparison across multiple LLMs, would help assess the robustness of our classifications. Lastly, there is an urgent need to enhance text analysis capabilities for African languages, allowing for a better representation of misinformation patterns associated with the specific cultural context of interest. However, given the paucity of published research analyzing geotagged social media posts from Sub-Saharan Africa, our study represents an important contribution.

It is highly likely that the influence of social media on the health information ecology in Sub-Saharan Africa will continue to grow [45]. It is also almost certain that future epidemics and pandemics will require rapid scale-up and public delivery of novel vaccines. Understanding how misinformation spreads in this context and how topics and tone both reflect and can predict public sentiment and attention will continue to be vital in the design of public health promotion campaigns and efforts to counter misinformation. Advancing methods for social media analysis applied in WEIRD contexts for the Low- and Middle-Income Countries (LMIC) context is crucial, and studies of this scale offer a foundation for that work to build on.

## Data Availability

Tweets cannot be shared publicly. We will provide Tweet IDs upon request.

## Acknowledgements

This study was partly supported by NIH-NIMHD (R01MD018340), the Penn Global Research Engagement Fund, and the Leonard Davis Institute of Health Economics. The funders had no role in study design, data collection and analysis, decision to publish, or preparation of the manuscript.

## Supporting information

### S1 Geolocation Validation and Data Cleaning

We used a geolocation algorithm to map tweets to countries from Sub-Saharan Africa. Then, we collected random samples of up to 3,000 tweets for each country from the mapped Sub-Saharan Africa set, and manually verified whether the self-reported location field corresponded to the assigned country. This step helped eliminate false positives due to ambiguous self-reported locations. Many high-volume countries (e.g., Kenya, Nigeria, South Africa, Uganda) exhibited zero false positives in the sampled data. In countries where false positives occurred, errors were driven by systematic and interpretable ambiguity patterns, such as homonymous place names or string collisions with U.S. state abbreviations (e.g., “WA,” which can refer to either Wa, a city in Ghana, or Washington State in the United States, and “Maryland,” which can refer to a U.S. state or to a county in Liberia), rather than by failures of the general mapping logic. Table S1 reports, for each country, the false positive (error) rate and representative examples of ambiguity patterns that led to misclassification. In total, **1,696,693** entries were removed from the geolocated dataset through this country-specific filtering process.

For the misinformation classification and LDA topic modeling pipelines, we preprocessed the corpus by mapping URLs and user mentions (@-mentions) to special tokens and removing duplicate tweets based on tweet content. The language of a tweet is automatically detected by the Twitter API (in ISO 639 language codes). In compliance with Twitter’s Terms of Service, we only include each tweet’s numeric tweet id (status ID, as given by the Twitter API) in the public release of the datasets.

**Table S1.** Country-level false positives identified during manual validation of geolocated tweets from Sub-Saharan Africa, based on random samples of up to 3,000 tweets per country. Representative user location strings causing misclassification are also provided.

| Country | False positive (Error) rate | Representative user location strings causing false positives |
| --- | --- | --- |
| Angola | 1.83% | “Zaire” |
| Benin | 0.40% | “Nikki” |
| Botswana | 0 | — |
| Burkina Faso | 0.17% | “Warsaw, Po”; “USA, Southwest”; non-African multi-location strings |
| Burundi | 0 | — |
| Cameroon | 0 | — |
| Cape Verde | 0 | — |
| Central African Republic | 2.03% | “Lafayette, Louisiana”; “NOLA”; “Atlanta, Chicago, Nola”; other U.S. city references |
| Chad | 0 | — |
| Comoros | 0 | — |
| Congo | 0 | — |
| Djibouti | 0 | — |
| Equatorial Guinea | 0 | — |
| Eritrea | 0 | — |
| Ethiopia | 0.17% | “Somali”; “Mogadishu, Somali” |
| Gabon | 0 | — |
| Gambia | 0.07% | “The Arsenal, North Bank” |
| Ghana | 83.1% | “WA”<br>(Washington State, United States) |
| Guinea | 0 | — |
| Guinea-Bissau | 0 | — |
| Ivory Coast | 19.8% | User locations containing the substring “, man” |
| Kenya | 0 | — |
| Lesotho | 0 | — |
| Liberia | 13.4% | User locations containing “, Maryland”<br>(United States) |
| Madagascar | 1.03% | “Bulgaria, Sofia”;<br>“Brussels, Luxembourg, Sofia”;<br>“Alaska, Madagascar”;<br>“Boston, Madagascar” |
| Malawi | 0 | — |
| Mali | 0 | — |
| Mauritania | 0 | — |
| Mauritius | 0 | — |
| Mozambique | 3.87% | “Palestine, Gaza” |
| Niger | 0 | — |
| Nigeria | 0 | — |
| Rwanda | 0 | — |
| São Tomé and Príncipe | 0 | — |
| Senegal | 0 | — |
| Sierra Leone | 0 | — |
| Somalia | 0 | — |
| South Africa | 0 | — |
| Tanzania | 0 | — |
| Togo | 0.47% | “Switzerland, Togo” |
| Uganda | 0 | — |
| Zambia | 0 | — |
| Zimbabwe | 0 | — |

### S2 Mapping Dictionaries

A mapping dictionary is used to look up location strings and map them to complete geographical locations. Each dictionary contains two values: *location string* and *mapped location*. The *location string* column is used to match self-reported location strings, and the *mapped location* column is the corresponding geographical location. For instance, if the location string is “Johannesburg, South Africa”, the corresponding mapping dictionary will have an entry with *location string* as “Johannesburg, South Africa” and *mapped location* as “Johannesburg, City of Johannesburg Metropolitan Municipality, Gauteng, South Africa”.

We create eleven such mapping dictionaries based on the number and type of administration levels (*AL*) present in a location string:

- *AL* = 1 has three dictionaries, containing either country, state, or city information in the *location string* column.
- *AL* = 2 has six dictionaries, containing information about one of the following pairs in the *location string* column: state-country, district-country, district-state, city-country, city-state, city-district.
- *AL* = 3 has two dictionaries, containing either distinct-state-country or city-state-country information in the *location string* column.

### S3 Stopwords

Prior to topic modeling, we removed two sets of stopwords. First, we removed the 100 most frequent terms in the corpus identified automatically by MALLET. Second, we removed a manually curated list of domain-specific stopwords consisting of retweet markers, URL and user placeholders, repeated punctuation, and highly frequent COVID-19-related keywords and hashtags.

#### Top 100 Most Frequent Terms Removed

*just, n, so, one, vaccination, what, on, by, people, about, how, %, they, more, ‘,’, rt, covid, from, covid-19, no, is, !, why, been, of, vaccine, now, after, against, when, out,…,:, i, we, there, at, to, was, us, nn, s, ‘’, [laughing emoji], are, should, the, in, if, will, get, \U0001f923, #covid19,., that, not, -, as, who, and,”,*

*<URL>, your, or, for, vaccines, be, with, up, it, (, this, my, vaccinated, first, ‘, an, our, do, all, have, t, pfizer, he, can,),*

*<USER>, la, health, a, you, has, ?, de, need, &, their, take, like, but*

#### Manually Curated Domain-Specific Stopwords

*RT, #pfizervaccine, #pfizercovidvaccine, #pfizerbiontech, #covid19vaccine, #moderna, #pfizer, #covid 19, #vaccinessavelives, #covidvaccine, #corona, #coronavaccine, #covidvaccines, #pandemic, #vaccination, coronavirus,…,…,….., rt, #covid, #coronavirus, #covid19, #covid-19, <URL>,*

*<URL>,*

*<USER>, covid, COVID, covid19, COVID19, COVID-19, covid-19, vaccine, vaccines, #vaccine, #vaccines, vaccination, vaccinations, moderna, pfizer, #coronavirusvaccine*.

### S4 Topic Modeling

We quantitatively assessed the quality of topics in each set by computing coherence scores using three measures of coherence, *C_V_* (0, 1), *C_UCI_* (0,1), and *C_NP MI_* (0,1), as well as topic uniqueness (diversity) (see Figure S1). We also reviewed the top words of topics within each topic set for a qualitative check of interpretability. Using both methods, we reached the consensus that the 100-topic model was optimal. Finally, we obtained the probability distribution for each tweet across all 100 topics.

**Note:** A small number of preprocessing artifacts (e.g., *→*, (*<*, and user*>*)) appeared among the top words; however, they occurred in less than 0.1% of tweets and therefore are unlikely to meaningfully affect the analyses.

**Fig S1.**
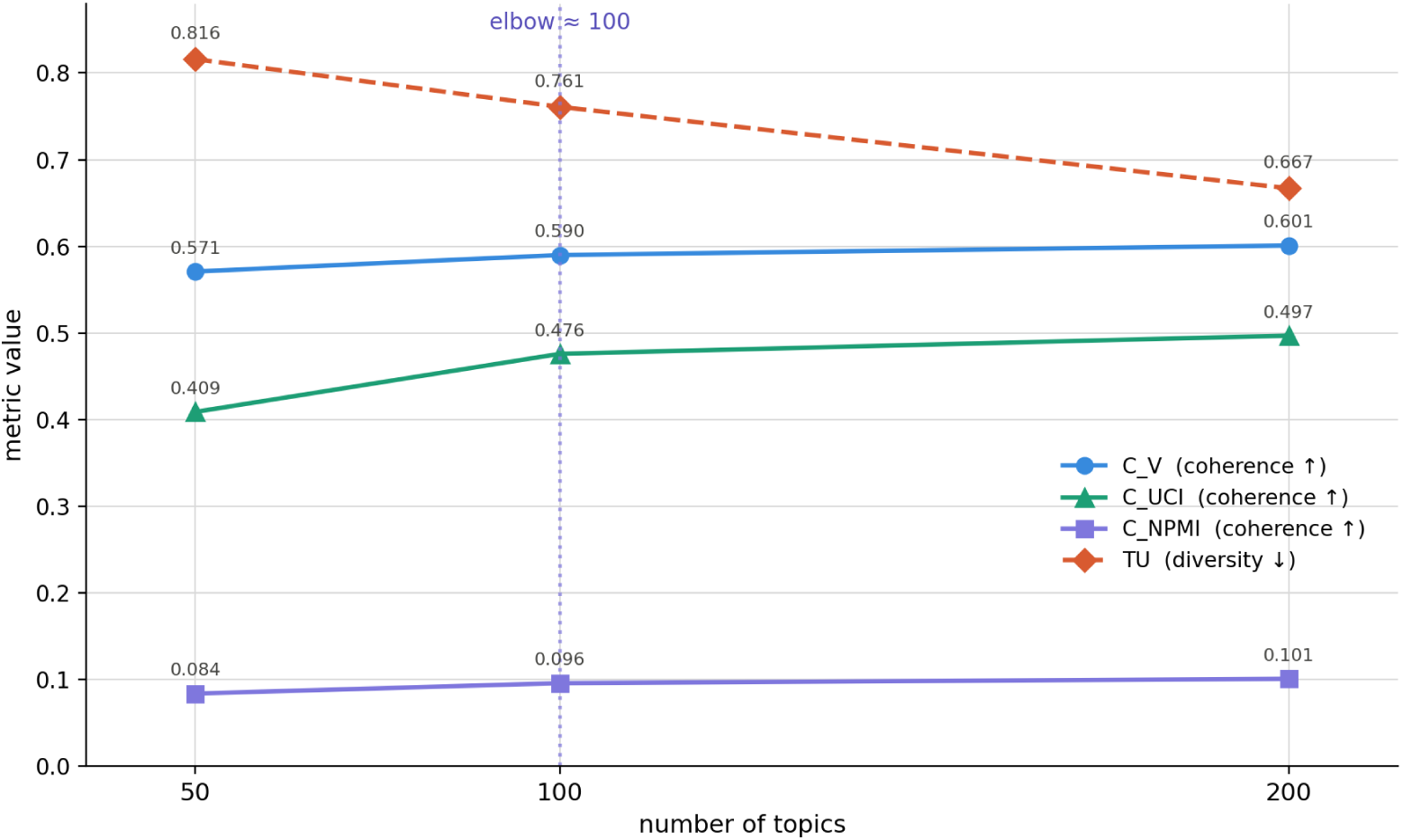
Topic model evaluation across LDA models with 50, 100, and 200 topics. Topic quality was assessed using three coherence metrics (*C_V_*, *C*_UCI_, and *C*_NPMI_; higher values indicate better coherence) and topic uniqueness (TU; lower values indicate less redundancy and greater topic distinctiveness). Coherence improved as the number of topics increased, whereas topic uniqueness declined. Based on the trade-off between coherence, diversity, and qualitative interpretability, the 100-topic model was selected for subsequent analyses.

### S5 Prompt for Misinformation Classification

We used GPT-4o-mini (temperature = 0) via the batch API to classify whether each tweet contained misinformation. The prompt used for classification is provided below:

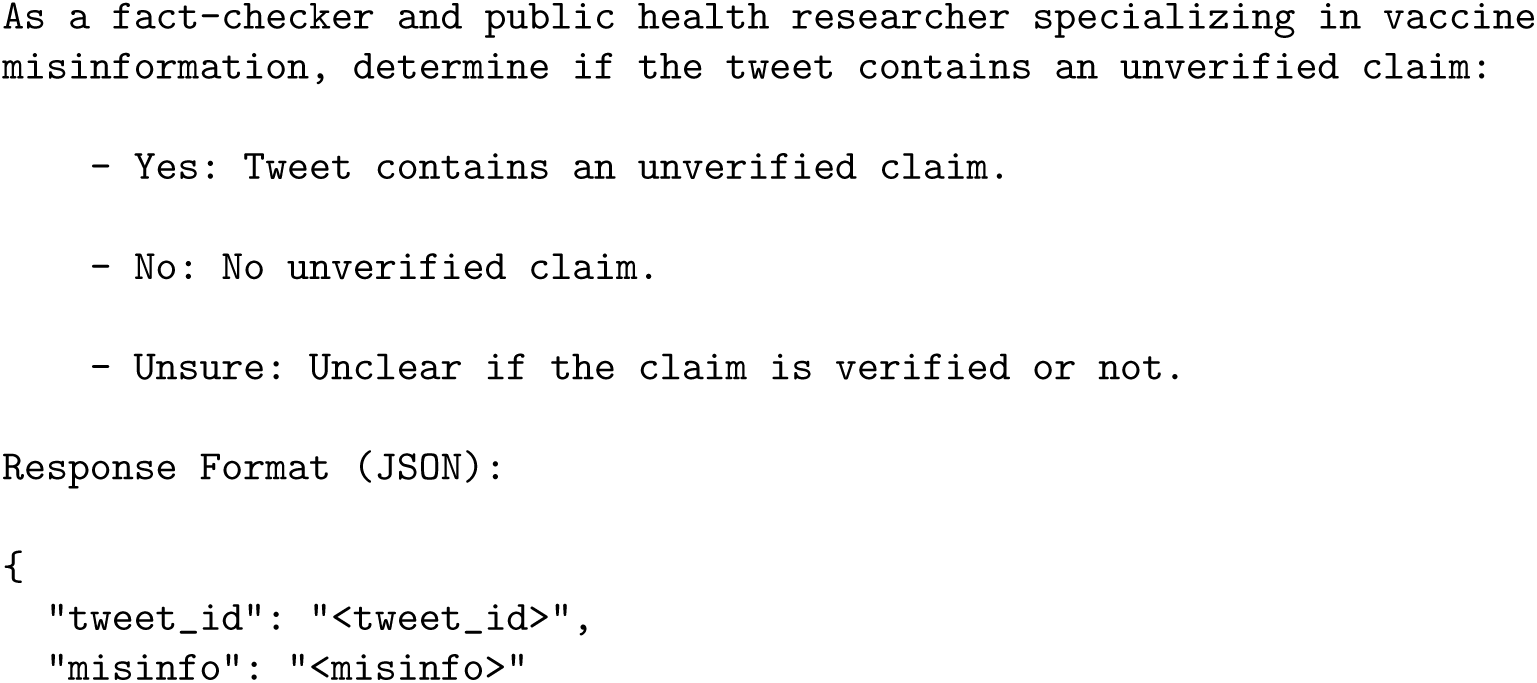
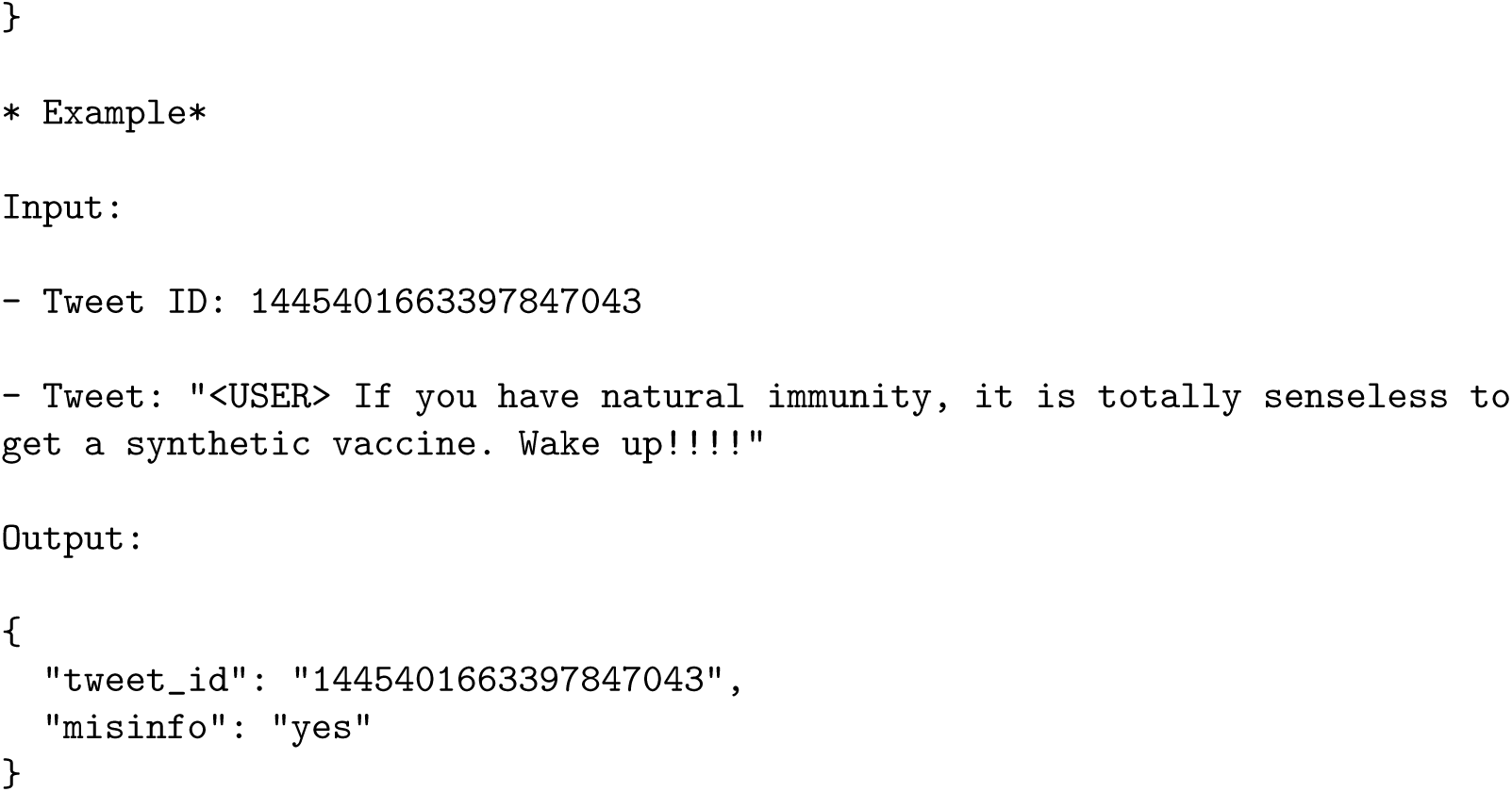

### S6 Descriptive Tables

**Table S2.**
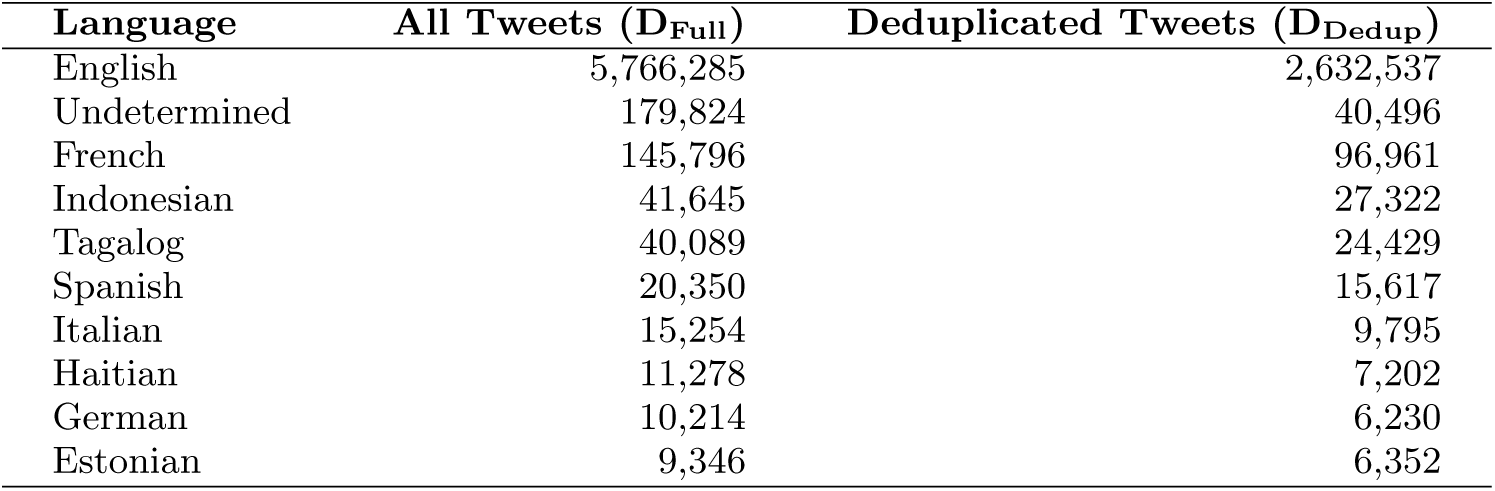
Top 10 language categories by frequency in the full geolocated (D_Full_) and deduplicated (D_Dedup_) Sub-Saharan Africa tweet datasets. Overall, D_Full_ contained 66 distinct language categories, and D_Dedup_ contained 65. Both datasets contained 61 identifiable languages, with English accounting for more than 90% of tweets. Twitter’s language detection algorithm labeled 179,824 tweets in D_Full_ and 40,496 tweets in D_Dedup_ as “undetermined.”

**Table S3.** Country-level descriptive statistics for the full (*D*_Full_) and deduplicated (*D*_Dedup_) Sub-Saharan Africa vaccine discourse datasets. For each country, the table reports its African region, income cluster, and numbers of tweets, unique users, and retweets. Region codes are 1: Central Africa, 2: Eastern Africa, 3: Southern Africa, and 4: Western Africa. Income codes are 1: Low income, 2: Lower-middle income, and 3: High/Upper-middle income. Countries are listed alphabetically.

| Country | Region | Income | $D_{Full}$ | | | $D_{Dedup}$ | | |
| --- | --- | --- | --- | --- | --- | --- | --- | --- |
|  |  |  | Tweets | Users | Retweets | Tweets | Users | Retweets |
| Angola | 1 | 2 | 7,571 | 1,168 | 5,007 | 5,154 | 820 | 3,009 |
| Benin | 4 | 1 | 4,903 | 601 | 3,899 | 2,782 | 428 | 1,875 |
| Botswana | 3 | 3 | 200,266 | 12,228 | 128,806 | 125,325 | 9,371 | 59,764 |
| Burkina Faso | 4 | 1 | 57,970 | 2,364 | 46,079 | 40,955 | 1,842 | 30,448 |
| Burundi | 2 | 1 | 3,832 | 646 | 2,563 | 2,381 | 491 | 1,201 |
| Cameroon | 1 | 2 | 88,070 | 9,290 | 57,978 | 56,024 | 6,966 | 28,713 |
| Cape Verde | 4 | 2 | 1,638 | 284 | 931 | 1,134 | 222 | 477 |
| Central African Republic | 1 | 1 | 1,227 | 183 | 764 | 847 | 137 | 412 |
| Chad | 1 | 1 | 2,998 | 314 | 1,522 | 2,198 | 263 | 791 |
| Comoros | 2 | 2 | 252 | 56 | 156 | 153 | 39 | 63 |
| Congo | 1 | 2 | 15,629 | 1,859 | 7,519 | 9,440 | 1,478 | 3,801 |
| Djibouti | 2 | 2 | 5,431 | 464 | 2,849 | 3,737 | 364 | 1,613 |
| Equatorial Guinea | 1 | 3 | 1,251 | 93 | 979 | 789 | 71 | 521 |
| Eritrea | 2 | 1 | 1,593 | 224 | 1,200 | 880 | 173 | 519 |
| Ethiopia | 2 | 1 | 39,932 | 5,672 | 29,807 | 16,877 | 3,363 | 9,257 |
| Gabon | 1 | 3 | 6,963 | 758 | 4,501 | 3,930 | 585 | 1,613 |
| Gambia | 4 | 1 | 4,732 | 743 | 3,294 | 2,778 | 555 | 1,451 |
| Ghana | 4 | 2 | 296,712 | 34,641 | 214,561 | 120,533 | 21,344 | 52,213 |
| Guinea | 4 | 1 | 3,296 | 323 | 2,550 | 2,348 | 243 | 1,674 |
| Guinea-Bissau | 4 | 1 | 52 | 23 | 33 | 29 | 18 | 12 |
| Ivory Coast | 4 | 2 | 18,225 | 2,378 | 13,632 | 10,761 | 1,538 | 6,644 |
| Kenya | 2 | 2 | 652,769 | 72,935 | 463,575 | 277,605 | 45,163 | 108,124 |
| Lesotho | 3 | 1 | 10,553 | 1,343 | 5,941 | 5,962 | 955 | 1,738 |
| Liberia | 4 | 1 | 3,755 | 584 | 2,125 | 2,352 | 464 | 891 |
| Madagascar | 2 | 1 | 14,654 | 1,000 | 11,455 | 9,073 | 780 | 6,114 |
| Malawi | 2 | 1 | 23,263 | 2,968 | 13,677 | 12,873 | 2,136 | 4,413 |
| Mali | 4 | 1 | 21,948 | 1,281 | 13,684 | 15,894 | 1,008 | 8,355 |
| Mauritania | 4 | 2 | 1,253 | 269 | 877 | 768 | 201 | 421 |
| Mauritius | 2 | 3 | 11,276 | 637 | 5,717 | 8,202 | 490 | 3,028 |
| Mozambique | 2 | 1 | 6,711 | 790 | 4,381 | 4,263 | 612 | 2,146 |
| Niger | 4 | 1 | 2,989 | 540 | 1,874 | 1,972 | 427 | 925 |
| Nigeria | 4 | 2 | 874,109 | 132,921 | 593,289 | 367,183 | 74,405 | 137,893 |
| Rwanda | 2 | 1 | 57,265 | 6,581 | 42,870 | 21,134 | 3,844 | 8,760 |
| Sao Tome and Principe | 1 | 2 | 18 | 1 | 10 | 8 | 1 | 1 |
| Senegal | 4 | 2 | 30,260 | 4,590 | 21,335 | 16,529 | 2,741 | 8,591 |
| Seychelles | 2 | 3 | 5,027 | 329 | 3,732 | 3,222 | 268 | 1,981 |
| Sierra Leone | 4 | 1 | 4,490 | 840 | 2,341 | 2,957 | 609 | 1,036 |
| Somalia | 2 | 1 | 21,958 | 2,563 | 15,386 | 13,847 | 1,868 | 7,879 |
| South Africa | 3 | 3 | 3,283,792 | 211,229 | 2,206,849 | 1,474,223 | 139,597 | 523,764 |
| Tanzania | 2 | 1 | 9,300 | 1,012 | 6,542 | 5,813 | 747 | 3,385 |
| Togo | 4 | 1 | 3,504 | 347 | 2,156 | 2,554 | 254 | 1,264 |
| Uganda | 2 | 1 | 309,750 | 34,080 | 216,545 | 142,233 | 23,806 | 56,465 |
| Zambia | 2 | 1 | 32,961 | 5,646 | 19,495 | 18,512 | 3,987 | 5,936 |
| Zimbabwe | 2 | 2 | 161,480 | 16,565 | 98,018 | 93,026 | 12,115 | 32,652 |
| **Total** | – | – | **6,305,628** | **573,363** | **4,280,504** | **2,909,260** | **366,789** | **1,131,833** |

**Table S4.**
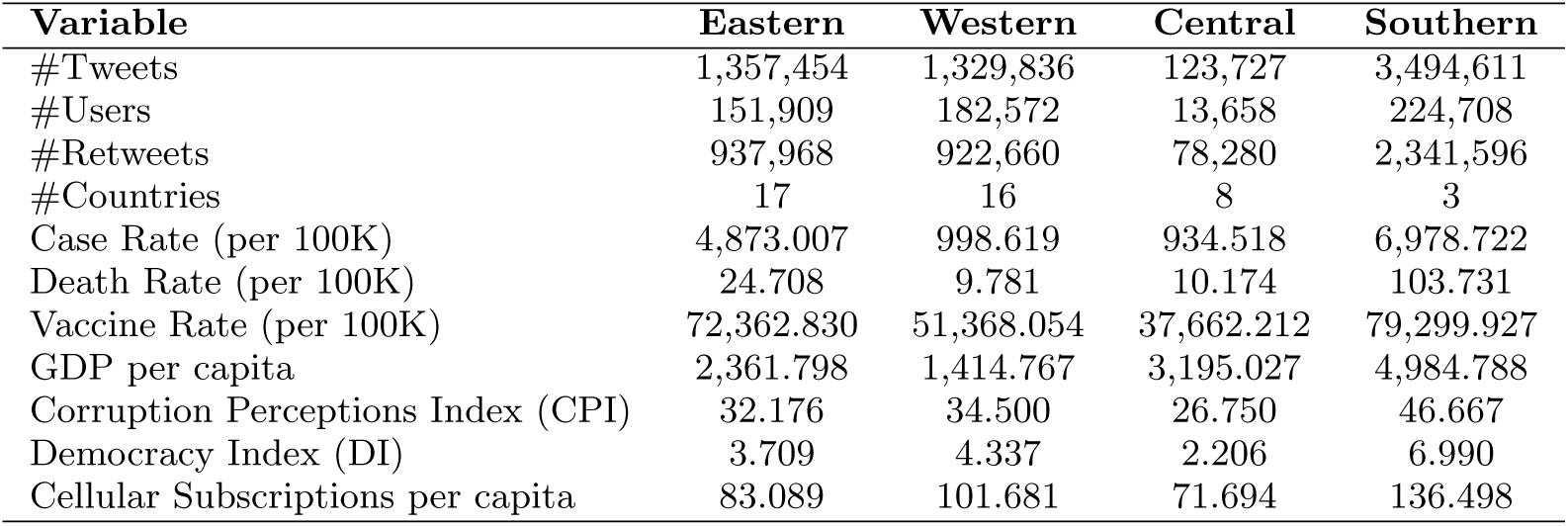
Descriptive statistics of Sub-Saharan African sub-regions and associated socioeconomic, governance, technological, and COVID-19 epidemiological indicators. COVID-19 case, death, and vaccination rates are reported per 100,000 population. Sub-regions comprise the following countries: *Eastern*: Burundi, Comoros, Djibouti, Eritrea, Ethiopia, Kenya, Madagascar, Malawi, Mauritius, Mozambique, Rwanda, Seychelles, Somalia, Tanzania, Uganda, Zambia, and Zimbabwe; *Western*: Benin, Burkina Faso, Cape Verde, Gambia, Ghana, Guinea, Guinea-Bissau, Ivory Coast, Liberia, Mali, Mauritania, Niger, Nigeria, Senegal, Sierra Leone, and Togo; *Central*: Angola, Cameroon, Central African Republic, Chad, Congo, Equatorial Guinea, Gabon, and S̃ao Tomé and Príncipe; *Southern*: Botswana, Lesotho, and South Africa.

| Variable | Eastern | Western | Central | Southern |
| --- | --- | --- | --- | --- |
| #Tweets | 1,357,454 | 1,329,836 | 123,727 | 3,494,611 |
| #Users | 151,909 | 182,572 | 13,658 | 224,708 |
| #Retweets | 937,968 | 922,660 | 78,280 | 2,341,596 |
| #Countries | 17 | 16 | 8 | 3 |
| Case Rate (per 100K) | 4,873.007 | 998.619 | 934.518 | 6,978.722 |
| Death Rate (per 100K) | 24.708 | 9.781 | 10.174 | 103.731 |
| Vaccine Rate (per 100K) | 72,362.830 | 51,368.054 | 37,662.212 | 79,299.927 |
| GDP per capita | 2,361.798 | 1,414.767 | 3,195.027 | 4,984.788 |
| Corruption Perceptions Index (CPI) | 32.176 | 34.500 | 26.750 | 46.667 |
| Democracy Index (DI) | 3.709 | 4.337 | 2.206 | 6.990 |
| Cellular Subscriptions per capita | 83.089 | 101.681 | 71.694 | 136.498 |

**Table S5.** Descriptive statistics of Sub-Saharan African country income clusters and their associated socioeconomic, governance, technological, and COVID-19 epidemiological characteristics. Countries were grouped into three income clusters: *Cluster 1 (Low income)*: Benin, Burkina Faso, Burundi, Central African Republic, Chad, Eritrea, Ethiopia, Gambia, Guinea, Guinea-Bissau, Lesotho, Liberia, Madagascar, Malawi, Mali, Mozambique, Niger, Rwanda, Sierra Leone, Somalia, Tanzania, Togo, Uganda, and Zambia; *Cluster 2 (Lower-middle income)*: Angola, Cameroon, Cape Verde, Comoros, Congo, Djibouti, Ghana, Ivory Coast, Kenya, Mauritania, Nigeria, S̃ao Tomé and Príncipe, Senegal, and Zimbabwe; *Cluster 3 (High/Upper-middle income)*: Botswana, Equatorial Guinea, Gabon, Mauritius, Seychelles, and South Africa. COVID-19 case, death, and vaccination rates are reported per 100,000 population.

| Variable | Cluster 1 | Cluster 2 | Cluster 3 |
| --- | --- | --- | --- |
| #Tweets | 643,636 | 2,153,417 | 3,508,575 |
| #Users | 70,578 | 277,219 | 225,203 |
| #Retweets | 450,183 | 1,479,737 | 2,350,584 |
| #Countries | 24 | 14 | 6 |
| Case Rate (per 100K) | 417.255 | 1,597.963 | 15,807.621 |
| Death Rate (per 100K) | 6.745 | 17.769 | 93.077 |
| Vaccine Rate (per 100K) | 44,343.347 | 61,633.401 | 110,691.081 |
| GDP per capita | 775.430 | 2,209.235 | 8,960.301 |
| Corruption Perceptions Index (CPI) | 29.917 | 33.286 | 44.833 |
| Democracy Index (DI) | 3.639 | 3.964 | 4.707 |
| Cellular Subscriptions per capita | 74.001 | 100.147 | 140.726 |

### S7 Topic Associations with Income Clusters

**Fig S2.**
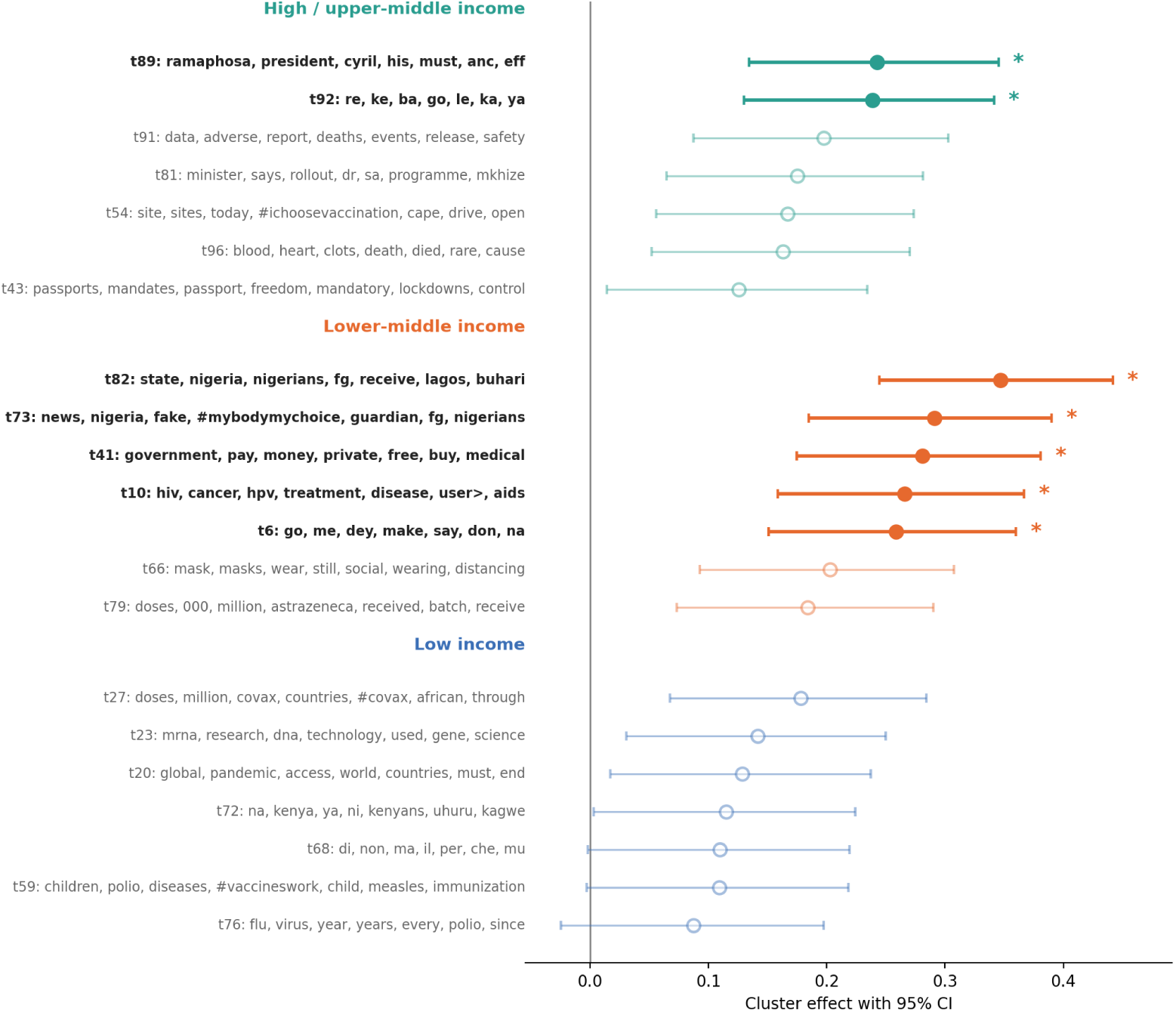
Topics associated with income clusters in Sub-Saharan Africa. Points represent standardized topic effects from differential language analysis, with horizontal bars indicating 95% confidence intervals. Values indicate topics overrepresented in a given income cluster. Filled markers (*) denote associations that remain significant after Benjamini-Hochberg correction. Income-related differences in vaccine discourse were limited, with only the lower-middle-income and high/upper-middle-income clusters showing significant overrepresentation of a small set of topics, while no robust topic associations emerged for the low-income clusters.

### S8 Regional Variation: Robustness Analysis

**Fig S3.**
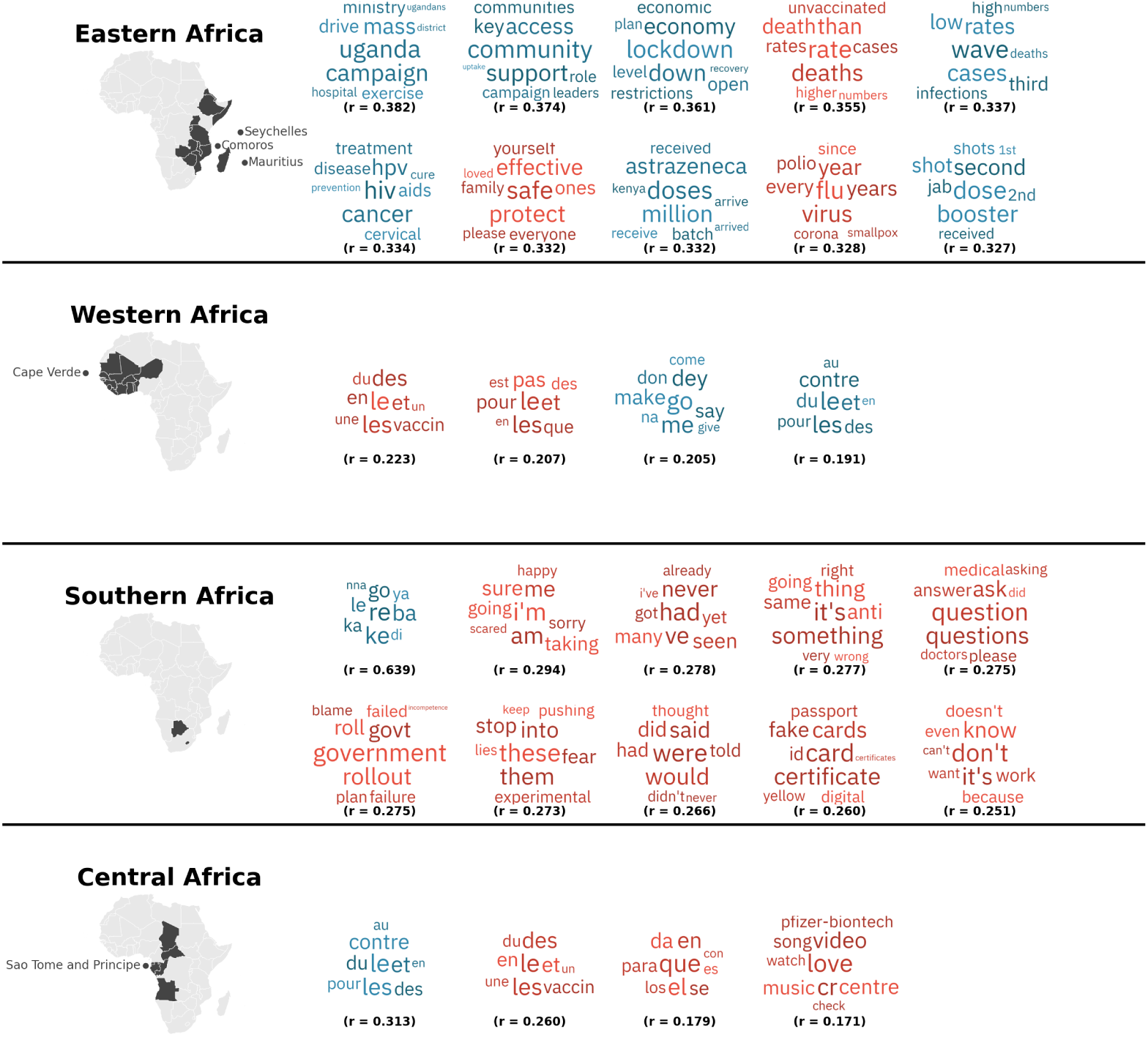
Regional variation in COVID-19 vaccination discourse across Sub-Saharan Africa after excluding each sub-region’s highest-volume country. For each African sub-region, the figure shows up to ten topics most strongly overrepresented relative to the other regions, following exclusion of South Africa, Nigeria, Kenya, and Cameroon — the highest-volume contributors to Southern, Western, Eastern, and Central Africa, respectively — as a robustness check on the regional characterizations reported in Figure 2. Topic cloud colors denote their relationship with misinformation discourse: **red** indicates topics positively associated with misinformation, **blue** indicates topics negatively associated with misinformation, and **gray** indicates no significant association. The correlation coefficients (*r*) shown below each topic cloud represent the strength of association between the topic and the focal region, computed on the reduced sample. Only topics significantly associated with a region after Benjamini-Hochberg correction (*p <* 0.05) are displayed. Eastern Africa’s implementation-focused characterization was largely retained after excluding Kenya, and Southern Africa’s emphasis on distrust and skepticism persisted after excluding South Africa. Western Africa’s expanded set retained the persisting non-misinformation Pidgin-conversational topic alongside three additional French-language topics. Central Africa’s topic set shifted slightly as well: the conspiracy-theory-specific topic present in the full sample did not persist after excluding Cameroon, suggesting it was disproportionately attributable to Cameroon rather than broadly characteristic of Central Africa.

## References

1. Moghadas SM, Vilches TN, Zhang K, Wells CR, Shoukat A, Singer BH, et al. The Impact of Vaccination on Coronavirus Disease 2019 (COVID-19) Outbreaks in the United States. Clinical Infectious Diseases. 2021 Dec 15;73(12):2257–64.

2. World Health Organization. WHO COVID-19 dashboard [cited 2026 Jul 6]. Available from: https://data.who.int/dashboards/covid19/vaccines

3. Dagovetz M, Momchilov K, Blank L, Khorsandi J, Rizzo A, Khabbache H, et al. Global COVID-19 vaccination challenges: Inequity of access and vaccine hesitancy. Journal of Medicine, Surgery, and Public Health. 2025 Aug 1;6:100197.

4. Wiysonge CS, Ndwandwe D, Ryan J, Jaca A, Batouré O, Anya BPM, et al. Vaccine hesitancy in the era of COVID-19: could lessons from the past help in divining the future?. Human vaccines & immunotherapeutics. 2022 Jan 31;18(1):1–3.

5. Kanyanda S, Markhof Y, Wollburg P, Zezza A. Acceptance of COVID-19 vaccines in sub-Saharan Africa: evidence from six national phone surveys. BMJ open. 2021 Dec 1;11(12):e055159.

6. de Figueiredo A, Temfack E, Tajudeen R, Larson HJ. Declining trends in vaccine confidence across sub-Saharan Africa: a large-scale cross-sectional modeling study. Human vaccines & immunotherapeutics. 2023 Jan 2;19(1):2213117.

7. Kigongo E, Kabunga A, Tumwesigye R, Musinguzi M, Izaruku R, Acup W. Prevalence and predictors of COVID-19 vaccination hesitancy among healthcare workers in Sub-Saharan Africa: A systematic review and meta-analysis. Plos one. 2023 Jul 28;18(7):e0289295.

8. Bangura JB, Xiao S, Qiu D, Ouyang F, Chen L. Barriers to childhood immunization in sub-Saharan Africa: A systematic review. BMC public health. 2020 Jul 14;20(1):1108.

9. Deml MJ, Githaiga JN. Determinants of COVID-19 vaccine hesitancy and uptake in sub-Saharan Africa: a scoping review. BMJ open. 2022 Nov 1;12(11):e066615.

10. Adamu AA, Essoh TA, Adeyanju GC, Jalo RI, Saleh Y, Aplogan A, et al. Drivers of hesitancy towards recommended childhood vaccines in African settings: a scoping review of literature from Kenya, Malawi and Ethiopia. Expert Review of Vaccines. 2021 May 4;20(5):611–21.

11. Ackah BB, Woo M, Stallwood L, Fazal ZA, Okpani A, Ukah UV, et al. COVID-19 vaccine hesitancy in Africa: a scoping review. Global health research and policy. 2022 Jul 19;7(1):21.

12. Unfried K, Priebe J. Vaccine hesitancy and trust in sub-Saharan Africa. Scientific Reports. 2024 May 13;14(1):10860.

13. Machingaidze S, Wiysonge CS. Understanding COVID-19 vaccine hesitancy. Nature medicine. 2021 Aug;27(8):1338–9.

14. Marcec R, Likic R. Using Twitter for sentiment analysis towards AstraZeneca/Oxford, Pfizer/BioNTech and Moderna COVID-19 vaccines. Postgraduate medical journal. 2022 Jul;98(1161):544–50.

15. Tasnim S, Hossain MM, Mazumder H. Impact of Rumors and Misinformation on COVID-19 in Social Media. Journal of preventive medicine and public health. 2020 Apr 2;53(3):171.

16. Pierri F, Perry BL, DeVerna MR, Yang KC, Flammini A, Menczer F, Bryden J. Online misinformation is linked to early COVID-19 vaccination hesitancy and refusal. Scientific reports. 2022 Apr 26;12(1):5966.

17. Piedrahita-Valdés H, Piedrahita-Castillo D, Bermejo-Higuera J, Guillem-Saiz P, Bermejo-Higuera JR, Guillem-Saiz J, et al. Vaccine Hesitancy on Social Media: Sentiment Analysis from June 2011 to April 2019. Vaccines. 2021 Jan;9(1):28.

18. Yan C, Law M, Nguyen S, Cheung J, Kong J. Comparing Public Sentiment Toward COVID-19 Vaccines Across Canadian Cities: Analysis of Comments on Reddit. Journal of medical Internet research. 2021 Sep 24;23(9):e32685.

19. Gabarron E, Oyeyemi SO, Wynn R. COVID-19-related misinformation on social media: a systematic review. Bulletin of the World Health Organization. 2021 Mar 19;99(6):455.

20. World Health Organization. Infodemic. [cited 2024 Mar 22]. Available from: https://www.who.int/health-topics/infodemic

21. Guntuku SC, Sherman G, Stokes DC, Agarwal AK, Seltzer E, Merchant RM, et al. Tracking Mental Health and Symptom Mentions on Twitter During COVID-19. Journal of general internal medicine. 2020 Sep;35(9):2798–800.

22. Kouzy R, Abi Jaoude J, Kraitem A, El Alam MB, Karam B, Adib E, Zarka J, Traboulsi C, Akl EW, Baddour K. Coronavirus goes viral: quantifying the COVID-19 misinformation epidemic on Twitter. Cureus. 2020 Mar 13;12(3).

23. Saha K, Torous J, Caine ED, Choudhury MD. Psychosocial Effects of the COVID-19 Pandemic: Large-scale Quasi-Experimental Study on Social Media. Journal of medical internet research. 2020 Nov 24;22(11):e22600.

24. Santosh R, Schwartz HA, Eichstaedt J, Ungar L, Guntuku SC. Detecting emerging symptoms of COVID-19 using context-based twitter embeddings. In Proceedings of the 1st Workshop on NLP for Covid-19 (Part 2) at EMNLP 2020 2020 Dec.

25. Skafle I, Nordahl-Hansen A, Quintana DS, Wynn R, Gabarron E. Misinformation About COVID-19 Vaccines on Social Media: Rapid Review. Journal of medical Internet research. 2022 Aug 4;24(8):e37367.

26. Alliheibi FM, Omar A, Al-Horais N. Opinion mining of Saudi responses to COVID-19 vaccines on Twitter. International Journal of Advanced Computer Science and Applications. 2021;12(6).

27. Boucher JC, Cornelson K, Benham JL, Fullerton MM, Tang T, Constantinescu C, et al. Analyzing Social Media to Explore the Attitudes and Behaviors Following the Announcement of Successful COVID-19 Vaccine Trials: Infodemiology Study. JMIR Infodemiology. 2021 Aug 12;1(1):e28800.

28. Kwok SWH, Vadde SK, Wang G. Tweet Topics and Sentiments Relating to COVID-19 Vaccination Among Australian Twitter Users: Machine Learning Analysis. Journal of medical Internet research. 2021 May 19;23(5):e26953.

29. Guntuku SC, Buttenheim AM, Sherman G, Merchant RM. Twitter discourse reveals geographical and temporal variation in concerns about COVID-19 vaccines in the United States. Vaccine. 2021 Jul 5;39(30):4034–8.

30. Herrera-Peco I, Jiménez-Gomez B, Romero Magdalena CS, Deudero JJ, García-Puente M, Benítez De Gracia E, et al. Antivaccine Movement and COVID-19 Negationism: A Content Analysis of Spanish-Written Messages on Twitter. Vaccines. 2021 Jun 15;9(6):656.

31. Offer-Westort M, Rosenzweig LR, Athey S. Battling the coronavirus ‘infodemic’ among social media users in Kenya and Nigeria. Nature Human Behaviour. 2024 May;8(5):823–34.

32. Tully M, Madrid-Morales D, Wasserman H, Gondwe G, Ireri K. Who is Responsible for Stopping the Spread of Misinformation? Examining Audience Perceptions of Responsibilities and Responses in Six Sub-Saharan African Countries. Digit Journal. 2022 May 28;10(5):679–97.

33. Blei DM, Ng AY, Jordan MI. Latent dirichlet allocation. Journal of machine Learning research. 2003;3(Jan):993–1022.

34. Schwartz HA, Giorgi S, Sap M, Crutchley P, Ungar L, Eichstaedt J. Dlatk: Differential language analysis toolkit. In Proceedings of the 2017 conference on empirical methods in natural language processing: System demonstrations 2017 Sep (pp. 55–60).

35. McCallum AK. MALLET: A Machine Learning for Language Toolkit. 2002; Available from: http://mallet.cs.umass.edu

36. Wanchoo K, Abrams M, Merchant RM, Ungar L, Guntuku SC. Reddit language indicates changes associated with diet, physical activity, substance use, and smoking during COVID-19. PLOS ONE. 2023 Feb 3;18(2):e0280337.

37. Röder M, Both A, Hinneburg A. Exploring the space of topic coherence measures. InProceedings of the eighth ACM international conference on Web search and data mining 2015 Feb 2 (pp. 399–408).

38. Mathieu E, Ritchie H, Rodés-Guirao L, Appel C, Gavrilov D, Giattino C, Hasell J, Macdonald B, Dattani S, Beltekian D, Ortiz-Ospina E. Excess mortality during the Coronavirus pandemic (COVID-19). Our world in data. 2020 May 22.

39. Johns Hopkins University & Medicine. Coronavirus Resource Center. [cited 2024 Mar 22]. Available from: https://coronavirus.jhu.edu/about/how-to-use-our-data

40. The World Bank. World Bank Open Data. [cited 2024 Mar 22]. Available from: https://data.worldbank.org

41. Zhao S, Hu S, Zhou X, Song S, Wang Q, Zheng H, et al. The prevalence, features, influencing factors, and solutions for COVID-19 vaccine misinformation: systematic review. JMIR public health and surveillance. 2023 Jan 11;9(1):e40201.

42. Wollburg P, Markhof Y, Kanyanda S, Zezza A. The evolution of COVID-19 vaccine hesitancy in Sub-Saharan Africa: evidence from panel survey data. InBMC proceedings 2023 Jul 6 (Vol. 17, No. Suppl 7, p. 8). London: BioMed Central.

43. Acheampong T, Akorsikumah EA, Osae-Kwapong J, Khalid M, Appiah A, Amuasi JH. Examining vaccine hesitancy in Sub-Saharan Africa: a survey of the knowledge and attitudes among adults to receive COVID-19 vaccines in Ghana. Vaccines. 2021 Jul 22;9(8):814.

44. Lasri K, Tonneau M, Naushan H, Malhotra N, Farouq I, Orozco-Olvera V, et al. Large-scale demographic inference of social media users in a low-resource scenario. In Proceedings of the International AAAI Conference on Web and Social Media 2023 Jun 2 (Vol. 17, pp. 519-529).

45. Ekezie W, Bosah G. Demographic representation of COVID-19 social media and information engagement in Nigeria. Population Medicine. 2021 Jun 28;3(June):1–6.

46. Encyclopaedia Britannica. Sub-Saharan Africa [Internet]. Encyclopaedia Britannica, Inc.; [cited 2026 Jul 31]. Available from: https://www.britannica.com/place/sub-Saharan-Africa

47. WorldAtlas. Regions of Africa [Internet]. WorldAtlas; [cited 2026 Jul 31]. Available from: https://www.worldatlas.com/geography/regions-of-africa.html

48. DLATK. TwitterMySQL [Internet]. GitHub; [cited 2026 Jul 31]. Available from: https://github.com/dlatk/TwitterMySQL

49. Jain D. Twitter-mapper [Internet]. GitHub; [cited 2026 Jul 31]. Available from: https://github.com/devanshrj/twitter-mapper

50. DLATK. [Internet]. DLATK Documentation; [cited 2026 Jul 6]. Available from: https://dlatk.github.io/dlatk/fwinterface/fwflag_lda_alpha.html

